# Sources of protein in UK adults following contemporary plant-based diets: findings from the Feeding the Future (FEED) Study

**DOI:** 10.64898/2026.11.23.26368579

**Authors:** Izabella Lawson, William Bell, Timothy J Key, Keren Papier

## Abstract

**Objective:** To identify and quantify sources of protein intake in United Kingdom (UK) adults following different diets, and assess how these sources contribute to meeting protein requirements.

**Design:** Cross-sectional analysis of the Feeding the Future Study (FEED), a UK cohort recruited in 2022-2023. Dietary intake was assessed using an online food frequency questionnaire. We calculated the percentage contribution of food groups to daily protein intake, and the proportion of participants meeting the estimated average requirement (EAR) for protein, with and without exclusion of under-reporters.

**Setting:** A contemporary cohort of UK adults.

**Participants:** 6,113 adults: 1,540 omnivores, 1,322 flexitarians, 547 pescatarians, 1,212 vegetarians, and 1,492 vegans.

**Results:** Grains contributed 10-30% of total protein intake across all groups. Dairy contributed 12.9-14.2% in non-vegan groups, while plant-based dairy alternatives contributed less in vegans (7%), mainly from soy milk (4.3%). Other major contributors (>10%) were meat for omnivores (33.0%) and flexitarians (13.5%); fish for pescatarians (14.9%); plant-based meat alternatives for vegetarians (12.0%); and plant-based meat alternatives (15.0%), legumes and pulses (10.5%), and nuts and seeds (10.5%) for vegans. The proportion below the protein EAR ranged from 1.9% in omnivores to 14.0% in vegans, falling to 0.01% and 3.6% respectively, after excluding under-reporters.

**Conclusions:** Grains were a major contributor to protein intake across all diet groups, as was dairy in all non-vegan groups. Protein sources were more varied in non-omnivorous groups, particularly vegans. Few participants fell below the protein EAR and the higher proportions among vegetarians and vegans were largely attenuated after excluding under-reporters.

## Introduction

The number of adults in the United Kingdom (UK) following plant-based diets has increased in recent years, with the proportion following a vegetarian or vegan diet doubling between 2008/2009 and 2018/2019 to reach around 3 million adults ^(1)^. Vegetarian diets exclude meat and fish, while vegan diets are devoid of all animal sourced foods, and both can include non-plant, non-animal foods such as fungi, seaweed, and yeast. The number of individuals reducing their meat intake is also increasing, in what is termed a flexitarian diet, where by 4% of UK adults now report only ‘occasionally’ eating meat ^(2)^.

Appropriately planned plant-based diets are generally considered nutritionally adequate, though this depends critically on food variety, life stage, and overall dietary composition ^(3)^. Plant-based diets have been associated with several health benefits including lower blood cholesterol levels, a lower body mass index (BMI), and lower risk of some of cardiometabolic diseases ^(4)^, likely related to their higher fibre content, lower energy density, and lower saturated fat content ^(4)^. Evidence from previous studies of UK adults following different plant-based diets suggests that, by restricting animal-source foods, plant-based diets may increase the risk of inadequate intakes of some nutrients, including protein ^(5, 6)^. These studies have shown that adults following vegan and vegetarian diets have, on average, lower protein intakes than those whose diets include meat and/or fish ^(7)^.

We observed similar findings in relation to total protein intake in the Feeding the Future (FEED) study, a contemporary cohort study of UK adults consuming different plant-based diets, despite the increased availability of plant-based dairy and meat alternatives in the food environment ^(8)^. Few studies, however, have characterised in detail the foods contributing to protein intake across contemporary UK diet groups, and further the implications for meeting population protein requirements. Understanding these differences will provide insight into protein adequacy, the key contributing protein sources, and additionally into the nutritional implications of these differences. Therefore this study aimed to first identify the key dietary sources of protein among adults following omnivorous, flexitarian, pescatarian, vegetarian, and vegan diets in the FEED study, and second, to assess the degree to which protein intake in plant-based dietary patterns are sufficient to meet population requirements.

## Methods

### Study population

FEED, an online study, was set up to collect up-to-date information on plant-based diets and diets containing meat and fish in the UK. Participants aged 18 or over were recruited between February 2022 and December 2023, and comprise ~6,700 adults’ resident in the UK. FEED participants were recruited through both paid and unpaid targeted and generic recruitment material via various UK institutions, societies and social media including the Vegan Society, the Vegetarian society, mainstream broadcasters such as radio stations and newsletters, and food distributors. Participation in the study required the completion of the baseline questionnaire that collected self-reported data on dietary motivations, anthropometrics, sociodemographic and lifestyle information, and dietary intakes. Further information on recruitment methods, the online questionnaire, and study rationale has been published elsewhere ^(5)^. The study was approved by the University of Oxford Medical Sciences Interdivisional Research Ethics Committee (MS IDREC) (R79226/RE002) on 22 January 2022.

### Dietary assessment

As part of the baseline questionnaire, participants were asked to select a diet group that best described their current diet and report on the length of time that they had been following the diet. Diet groups were self-reported and selected from the following options: omnivore (consume meat frequently), flexitarian (infrequent meat eating), pescatarian (consume fish but no meat), vegetarian (do not consume meat or fish), and vegan (do not consume any animal-source foods). Usual dietary intake over the previous 12 months was assessed using the FEED food frequency questionnaire (FFQ). This FFQ was adapted from the validated FFQ used by the UK centres participating in the European Prospective Investigation into Cancer and Nutrition (EPIC) ^(9–11)^, and modified to include a wider range of plant-based meat, dairy and egg alternatives with questions to determine types, brands, and fortification specifications of any plant-based alternatives consumed. Individual and composite foods in the FFQ were assigned nutrient values based on foods and recipes included in the UK Nutrient Data Bank (NDB) or from product manufacturer data where available. We estimated daily food and beverage intakes (in g or ml) using standard portion sizes as used in the Oxford WebQ dietary recall questionnaire ^(12)^. When Oxford WebQ data were not available we used information from UK Ministry of Agriculture, Fisheries and Food data ^(13)^ and product manufacturers’ websites. Daily individual intakes of nutrients were calculated using the UK NDB extract version: UK_NDB_1.2, 2023 ^(14)^, and FoodDB (up to May 2022) ^(15)^ cross-checked against online retailers for plant-based alternatives not incorporated in the NDB ^(5)^.

### Exclusions

Of the 6,684 FEED participants recruited, 246 participants were excluded due to incomplete questionnaire data, and 325 were excluded due to implausible dietary intakes (defined as <500 kcal/day(d) and >3500 kcal/d for women and <800 kcal/d and >4000 kcal/d for men) or where more than 20% of relevant FFQ items for each diet group were missing ^(16)^; after these exclusions the analysis sample was 6,113 participants.

### Statistical analysis

Descriptive statistics were used to summarise participant characteristics and mean macronutrient intakes by diet group; results are presented as means and standard deviations (SD) or as counts and percentages. Significant between group differences were tested for using ANOVA or Chi-square tests as appropriate; for nutrient intakes, pairwise comparisons were conducted using Wald’s t-tests, adjusted for multiple comparisons using a Bonferroni correction. Survey food items were classified into 16 primary food groups, and also categorised into three broad categories: exclusively animal-source foods, exclusively plant-source foods, and mixed (containing both animal and plant-derived foods) (Supplementary Table 1). We calculated age- and gender-adjusted mean daily intakes of all foods and beverages (in g or ml) for each dietary group using multivariable linear regression, and derived adjusted mean values and standard errors using the least squares mean statistic via the emmeans package in R ^(5)^. To assess the contribution of individual food items to total protein intake, the protein provided by each item (g) was divided by the total protein intake (g) within the corresponding dietary group. The protein contribution of each food group was determined by calculating the proportion of protein provided by all items belonging to the food group relative to the total protein intake from all foods and beverages consumed within each diet group. All statistical analyses were performed using Stata version 18.5 and R version 4.1.

### Sensitivity Analysis

We previously found that 10-15% of vegetarians and vegans in the FEED study had estimated protein intakes categorised as inadequate ^(5)^, based on the Estimated Average Requirement (EAR; ≥0.66 g/kg body weight) ^(17)^. Global underreporting of dietary intake may result in the misclassification of participants as having inadequate protein intakes. Thus, we conducted a sensitivity analysis excluding potential under-reporters according to the Goldberg cut-off method, which is based on the ratio of reported energy intake-to-basal metabolic rate (EI:BMR) <1.2 ^(18)^. We recalculated the proportion of those meeting the EAR for protein and repeated all analyses after excluding 1,413 participants with an EI:BMR ratio <1.2 ^(19)^ or with missing body weight data, resulting in a sensitivity analysis sample of 4,700 participants. BMR was calculated using the Schofield equation ^(20)^.

## Results

Table 1 shows the mean daily energy and macronutrient intakes by diet group adjusted for age and gender. The large majority of participants were women. Age and BMI differed across the diet groups (P heterogeneity <0.001). Vegans were youngest on average (mean ± SD: 24.3 ± 4.8 years) and pescatarians were oldest on average (mean ± SD: 53.1 ± 4.8 years), while flexitarians and vegans had the lowest mean BMI and omnivores the highest (mean ± SD: flexitarians = 24.1 ± 4.9 kg/m^2^; vegans = 24.1 ± 4.9 kg/m^2^, omnivores = 26.2 ± 5.8 kg/m^2^). A similar pattern was observed across genders. Protein intake was significantly different across diet groups (P heterogeneity <0.001) and decreased with greater animal food exclusion both in terms of total intake and percent energy (mean ± SD: 104.4 ± 35.3 g/d or 17.8 ± 4.5%E in omnivores vs 69.1 ± 23.3 g/d or 12.2 ± 2.3%E in vegans). In pairwise comparisons, protein intakes in all diet groups were significantly different from each other with the exception of vegan and vegetarian diets. Total energy intake was also significantly different across the diet groups (P heterogeneity <0.001) and was highest among omnivores (mean ± SD: 2268 ± 577 kcal/d) and flexitarians (mean ± SD: 2237 ± 604 kcal/d) and lowest among vegans (mean ± SD: 2172 ± 604 kcal/d) and vegetarians (mean ± SD: 2154 ± 574 kcal/d), with significant differences observed between omnivores and vegetarians and vegans, and flexitarians and vegetarians and vegans. The proportion of participants categorised as having protein intakes below the EAR was significantly different across diet groups (P heterogeneity <0.001) and increased as animal foods were excluded from the diet, ranging from 1.9% of omnivores to 14.0% of vegans. In pairwise comparisons, all diet groups differed significantly from each other with the exception of omnivores and flexitarians, and vegetarians and vegans.

**Table 1.** Participant characteristics and daily nutrient intakes by diet group in 6,113 FEED participants.

|  | **Overall** |  | **Omnivore** |  | **Flexitarian** |  | **Pescatarian** |  | **Vegetarian** |  | **Vegan** |  |  |
|---|---|---|---|---|---|---|---|---|---|---|---|---|---|
| **N** | 6,113 |  | 1,540 |  | 1,322 |  | 547 |  | 1,212 |  | 1,492 |  | ***P***<sup>‡</sup> |
| % n | 100% |  | 25.2% |  | 21.6% |  | 8.9% |  | 19.8% |  | 24.4% |  |  |
| **Characteristics** | **N** | **%** | **N** | **%** | **N** | **%** | **N** | **%** | **N** | **%** | **N** | **%** |  |
| Gender |  |  |  |  |  |  |  |  |  |  |  |  | <0.001 |
| Women | 4,931 | 80.7 | 1225 | 79.5% | 1128 | 85.3% | 480 | 87.8% | 996 | 82.2% | 1102 | 73.9% |  |
| Men | 1,084 | 17.7 | 306 | 19.9% | 174 | 13.2% | 64 | 11.7% | 200 | 16.5% | 340 | 22.8% |  |
| Prefer not to say | 98 | 1.6 | 9 | 0.6% | 20 | 1.5% | - | - | 16 | 1.3% | 50 | 3.4% |  |
|  | **Mean** | **SD** | **Mean** | **SD** | **Mean** | **SD** | **Mean** | **SD** | **Mean** | **SD** | **Mean** | **SD** |  |
| Age (years) | 51.2 | 15.9 | 52.7 | 14.6 | 51.9 | 16.9 | 53.1 | 16.3 | 52.3 | 16.4 | 47.5 | 15.2 | <0.001 |
| BMI (kg/m<sup>2</sup>) | 24.9 | 5.3 | 26.2 | 5.8 | 24.1 | 4.9 | 24.4 | 5.1 | 24.9 | 5.4 | 24.3 | 4.8 | <0.001 |
| Women | 25.2 | 4.4 | 26.2 | 6.0 | 24.0 | 5.0 | 24.3 | 5.2 | 24.8 | 5.5 | 24.2 | 5.1 |  |
| Men | 24.8 | 5.5 | 26.2 | 4.7 | 24.9 | 4.5 | 24.7 | 3.7 | 25.5 | 4.8 | 24.3 | 3.7 |  |
| Prefer not to say | 24.9 | 5.7 | 27.5 | 7.3 | 23.7 | 4.6 | - | - | 25.0 | 6.0 | 24.3 | 5.3 |  |
| **Mean Nutrient Intakes**<sup>†</sup> |  |  |  |  |  |  |  |  |  |  |  |  |  |
|  | **Mean** | **SD** | **Mean** | **SD** | **Mean** | **SD** | **Mean** | **SD** | **Mean** | **SD** | **Mean** | **SD** |  |
| Protein (g/day) | 83.5 | 29.9 | 104.4 | 35.3 | 87.8 | 23.3 | 81.6 | 24.2 | 70.8<sup>a</sup> | 20.9 | 69.1<sup>a</sup> | 23.3 |  |
| Energy (kJ) | 9284 | 2447 | 9515<sup>b</sup> | 2423 | 9398<sup>b</sup> | 2380 | 9291<sup>a,b</sup> | 2442 | 9051<sup>a</sup> | 2408 | 9130<sup>a</sup> | 2537 |  |
| Energy (kcal) | 2210 | 583 | 2268<sup>b</sup> | 577 | 2237<sup>b</sup> | 567 | 2211<sup>a,b</sup> | 582 | 2154<sup>a</sup> | 574 | 2172<sup>a</sup> | 604 |  |
| Fat (%E) | 36.7 | 8.3 | 40.3 | 11.2 | 36.3<sup>b</sup> | 6.4 | 35.4<sup>a,b</sup> | 7.0 | 34.9<sup>a</sup> | 6.3 | 35.2<sup>a</sup> | 6.7 |  |
| Carbohydrate (%E) | 45.1 | 10.2 | 38.0 | 13.8 | 44.8 | 6.9 | 46.4 | 7.2 | 48.8<sup>a</sup> | 6.7 | 49.1<sup>a</sup> | 6.9 |  |
| Alcohol (%E) | 2.7 | 3.5 | 2.8<sup>a</sup> | 3.9 | 2.8<sup>a</sup> | 3.2 | 3.1<sup>a</sup> | 3.7 | 2.7<sup>a</sup> | 3.3 | 2.2 | 3.2 |  |
| Protein (%E) | 14.5 | 3.7 | 17.8 | 4.5 | 15.1 | 2.5 | 14.1 | 2.3 | 12.6 | 2.0 | 12.2 | 2.3 |  |
|  | **N** | **%** | **N** | **%** | **N** | **%** | **N** | **%** | **N** | **%** | **N** | **%** |  |
| **Proportion at or above Protein EAR** | 5,415 | 92.6 | 1461 | 98.1% | 1241 | 96.7% | 491 | 94.2%<sup>b</sup> | 1010 | 88.6%<sup>a</sup> | 1212 | 86.0%<sup>a</sup> | <0.001 |

BMI = Body Mass Index; %E = Percent energy, EAR = Estimated Average Requirement
BMI excludes 268 participants with missing bodyweight data. Age and mean nutrient intakes exclude 1 participant with missing age.
a,b Estimates sharing a letter are not significantly different (p ≥ 0.05), based on pairwise comparisons of adjusted means following ANOVA with Bonferroni correction
All mean nutrient intakes P heterogeneity <0.001
<sup>†</sup>Mean intakes adjusted for age and gender
<sup>‡</sup>P values for ANOVA and Chi-square tests
Data not shown where categories have <5 participants

The categorisation of individual FFQ items into food groups and the proportion contribution to total protein by broad categories (animal-source foods, plant-source foods, and mixed) by diet group is presented in Figure 1 and Supplementary Table 1. The majority of total protein intake in omnivores came from animal sources (63%), for flexitarians the contribution from animal and plant protein sources was approximately even (44% and 47% respectively), and the majority of protein for pescatarians (57%), vegetarians (68%), and vegans (100% including mixed categories) came from plant-based protein sources. Across all diets, grains consistently emerged as a top contributor to protein intake, as did dairy in non-vegan groups (>10% contribution to total protein intake), while other food groups such as, fish, legumes, and meat (or plant-based alternatives) varied according to dietary exclusions. Among omnivores, food groups contributing more than 10% to total protein intake included primarily meat (33.0%), grains (13.5), and fish (11.2%). In flexitarians, grains (19.4%), meat (13.4%), and fish (12.5%). In pescatarians; grains (21.4%), and fish (14.9%). In vegetarians; grains (25.8%), and plant-based meat alternatives (12.0%). In vegans, grains (27.2%), plant-based meat alternatives (15.0%), vegetables (including peas and green beans) (11.0%), nuts and seeds (10.5%) and pulses and legumes (10.5%).

**Figure 1:**
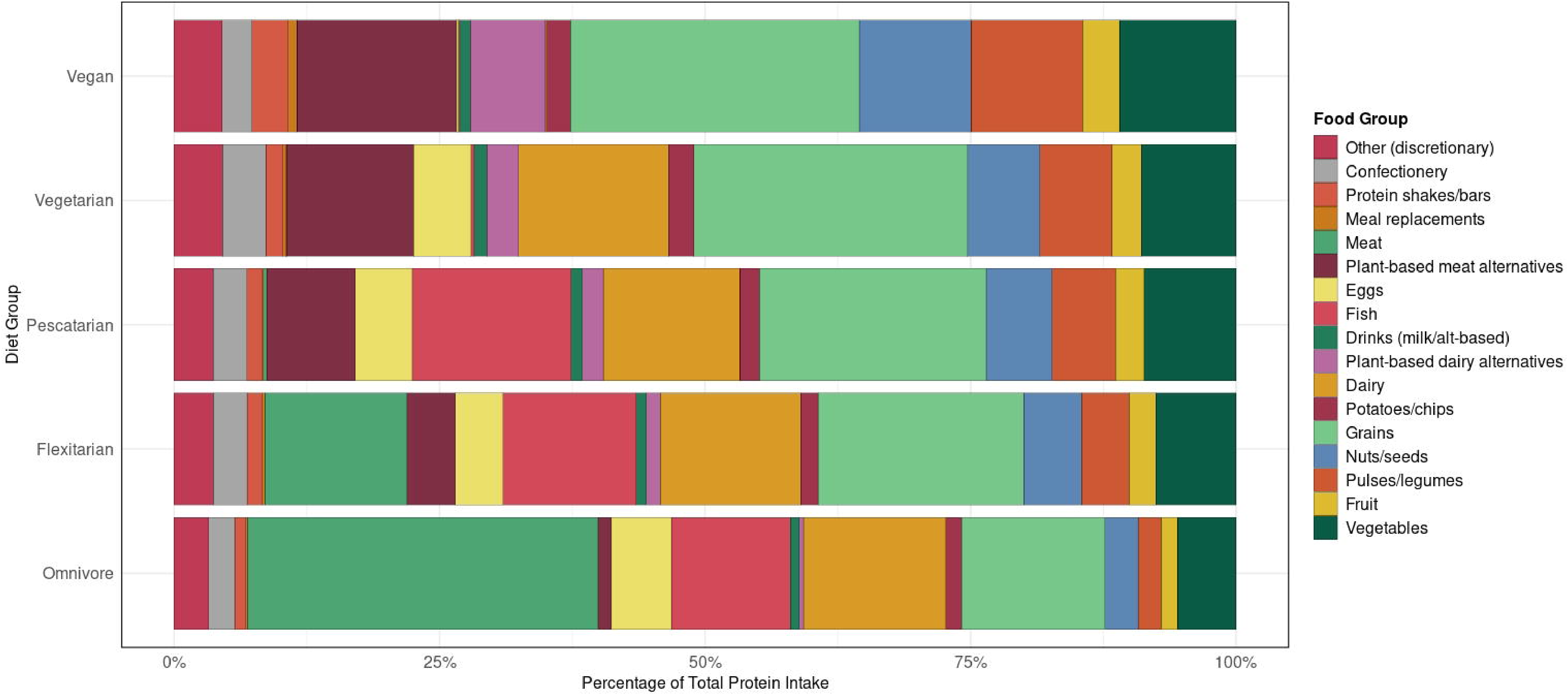
Protein contribution (%) by food group and diet in 6,113 FEED participants.

The top 10 contributing foods to total protein intake in each diet group are shown in Figure 2, and the age and gender adjusted mean daily intakes for all foods and beverages consumed and their respective individual contribution to total protein by diet group are shown in Supplementary Table 2. For omnivores, poultry and beef were the top contributors making up 8.7% and 8.6% of total protein intake respectively. In other diet groups, the relative contribution of the top 10 food sources to total protein intake were lower. For flexitarians, poultry was the top individual contributor (4.6%), closely followed by eggs (4.5%) and oily fish (4.3%). The top individual sources of protein among pescatarians were eggs (5.4%) and oily fish (5.1%). Individually, the top sources for vegetarians were eggs (5.4%), cheese (5.2%), and wholemeal bread (5.1%). The top individual sources of protein in vegans were wholemeal bread (5.5%), followed by nuts (5.0%), pulses (4.4%) and soy milk (4.3%).

**Figure 2:**
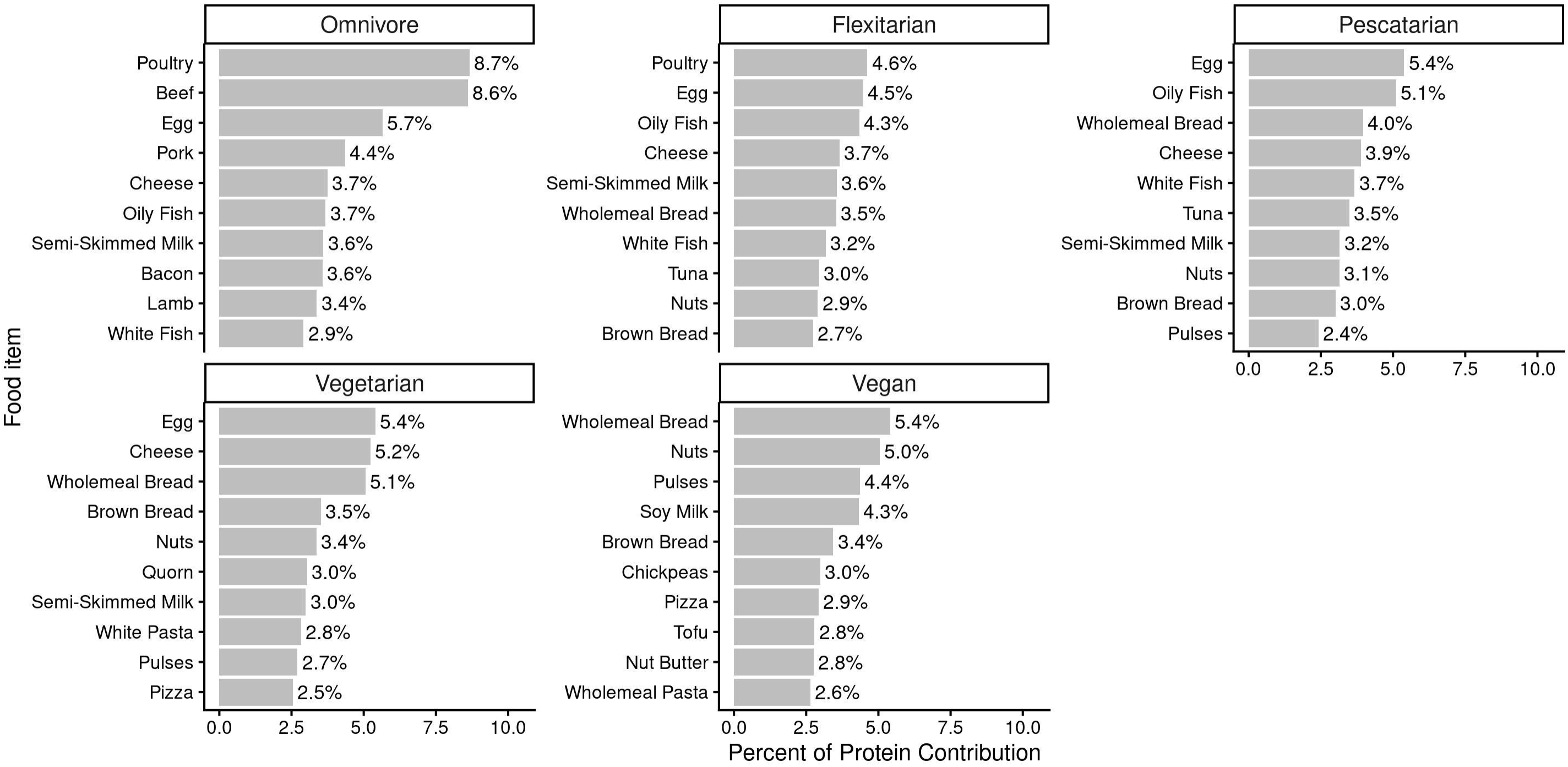
Top 10 individual percentage protein contributors by diet group ranked in 6,113 FEED participants.

### Sensitivity analysis

Supplementary Table 3 presents the mean daily total energy and macronutrient intakes by diet groups adjusted for age and gender after additionally excluding participants with EI:BMR ratio <1.2. Concurrent with the full analysis sample, age and BMI differed significantly across the diet groups, whereby vegans were youngest on average (mean ± SD: 48.9 ±15.2) and pescatarians were oldest (mean ± SD: 54.5 years). Flexitarians and vegans had the lowest BMI on average (mean ± SD: flexitarians = 23.7 ± 4.3kg/m^2^, vegans = 23.7 ± 4.4 kg/m^2^), and omnivores the highest (mean ± SD: 25.6 ± 5.1). The pattern of nutrient intakes remained largely unchanged; significant differences were observed across diet groups for protein, energy, and the proportion of those meeting the EAR (P heterogeneity <0.001); protein intakes were lower in diets with greater animal food exclusion both in terms of total intake and percent energy (mean ± SD: 110.7 ± 35.0 g/day or 17.4 ± 4.3%E in omnivores vs 74.7 ±21.7 g/day or 12.0 ±2.3%E in vegans), with significant differences observed between all diet groups except vegetarians and vegans. Energy intakes were however higher in the sensitivity analysis sample than the overall sample across all diet groups, and were highest in the omnivores (mean ± SD: 2435 ± 485 kcal/d) and pescatarians (mean ± SD: 2396 ± 493 kcal/d) and lowest among vegans (mean ± SD: 2363 ± 497 kcal/d) and vegetarians (mean ± SD: 2339 ± 514 kcal/d), while significant differences were observed between all groups except omnivores and flexitarians and vegetarians and vegans (Supplementary Table 3). The proportion of participants below the protein EAR was substantially reduced, ranging from <0.05% in omnivores, flexitarians, and pescatarians, to 2% in vegetarians, and 3.6% in vegans. Age and gender adjusted macronutrient intakes in the subsample were also higher than in the full analysis sample, while the pattern of individual food contributions to total protein intake in the sensitivity analysis sample was consistent with the full analysis sample (Supplementary Table 4).

## Discussion

This study describes the major dietary sources of protein in five distinct diet groups within the FEED cohort and further investigates the degree to which protein intake across the diet groups is sufficient to meet population requirements. Specifically, grains consistently represented a major source of protein across all diet groups, as did dairy in all non-vegan groups. Among meat eaters (omnivores and flexitarians), animal-source food groups such as meat and fish were also dominant protein sources, whereas in pescatarians, fish provided the majority of protein alongside dairy and grains. Among vegetarians, protein intake was largely derived from dairy and plant-based meat alternatives in addition to grains. Vegans relied on a wide range of plant-source food groups for their protein intake, including plant-based meat alternatives, vegetables, nuts and seeds, pulses and legumes, and grains. Further breakdown of specific food contributors highlighted that meat (particularly poultry and beef) was a key source of protein for omnivores, while eggs and fish were top individual sources of protein for flexitarians and pescatarians, respectively. Eggs, cheese, and bread were the top individual protein contributors in vegetarians, whereas vegans relied on bread, nuts, soy milk, pulses and legumes. Total protein intake decreased with greater animal food exclusion, as did the proportion of participants not meeting the EAR for total protein intake, with significant pairwise differences between all groups except vegetarians and vegans. In a sensitivity analysis removing under-reporters, the proportion of participants with protein intakes below the EAR decreased, particularly in groups that relied more heavily on plant-based protein sources.

Several studies, including our previously published findings from the FEED cohort, have reported higher consumption of a wide variety of plant-based foods in low and non-meat-eaters ^(5, 21-24)^. A review of 13 observational studies conducted across the United States, Europe (including the UK), and South Korea reporting on protein intake and sources in vegan, vegetarian, and semi-vegetarian diets found that vegans exhibited the greatest diversity in their intake of plant-based protein sources and demonstrated the highest reliance on cereals compared to other diet groups ^(25)^. In the FEED cohort, the diversity was even more pronounced: although grains were an important contributor, they accounted for roughly a quarter of total protein in vegan diets, in contrast to 60.3% reported in the review. It is important to note, however, that the definition of cereals and grains in other studies was broader and would also encompass food items that were categorised separately in our analysis (e.g. confectionery, discretionary foods). The diets of flexitarians are less well documented in the previous literature. Studies often report on ‘semi-vegetarians’, characterised by a lower, but not totally absent meat intake. Protein intake of ‘semi-vegetarians’ in previous studies ranges from 13.7% to 16.5% of total energy, with the proportion of total protein intake coming from plant sources varying substantially from 37% to 83% ^(25)^. The proportion of total energy from protein in FEED was 15.1%, and comparatively flexitarians (87.8g/day) had a lower total protein intake than omnivores (104.4g/day), but higher than pescatarians (81.6g/day), vegetarians (70.8g/day), and vegans (69.1g/day). Flexitarians still consumed a large variety of animal-based protein sources, though the sources varied in comparison to omnivores, with flexitarians consuming less red and processed meat, opting for poultry, fish, eggs, and dairy instead. Interestingly, the main dietary sources of protein reflected more closely to that of vegetarians and pescatarians than omnivores. Previous literature also highlights limitations in capturing the intake and composition of plant-based meat and dairy substitutes, making it challenging to assess their contribution to overall macronutrient and micronutrient intakes ^(25)^. Although vegetarians and vegans in our cohort relied primarily on other sources of protein such as grains and legumes as the main individual sources of protein, plant-based meat and dairy alternatives emerged as one of the top protein sources when contribution was examined at the food group level, although compared to plant-based meat alternatives, plant-based dairy alternatives contributed less in vegetarians compared to vegans. Similar findings have been reported in a sample of Norwegian vegans, where 19% of protein was provided by meat and dairy substitutes ^(26)^, suggesting that plant-based meat and dairy alternatives may play an increasingly important role in supplying protein in contemporary diets limiting or excluding animal sourced foods.

The lower total protein intake in vegans and vegetarians in FEED is consistent with previous literature including in UK cohorts ^(7, 25, 27)^. Notably, however protein intake in vegans was only marginally lower than vegetarians and not significantly different after adjustment for age and gender. National Diet and Nutrition Survey (NDNS) data show, in adults aged between 18-64 years in the UK, the mean daily protein intake is 70.2g/day or 16.9% of total energy. Omnivores in FEED show a similar protein intake when considered as a % of total energy intake, although total protein intake in g is higher overall at 104.4g/day. Notably, vegans and vegetarians show a similar absolute intake of approximately 70.0g/day, but this is lower when considering it as a % of total energy intake, while flexitarians and pescatarians consume more than the population average in g/day, but slightly less when considering protein as a % of total energy. These differences may reflect the difference in dietary assessment methods between the studies, or differences in demographics and the study population such as the inclusion of older adults on average in FEED. Overall, in the total population around seven percent of FEED participants had protein intakes below the EAR, and the proportion of individuals below the EAR within each diet group increased as the consumption of animal-source foods decreased, with nearly 14% of vegans below the EAR. Dietary under-reporting was prevalent across all diet groups, although rates were slightly higher in vegetarians and vegans compared to other groups. A sensitivity analysis excluding the 1,461 participants with implausibly low energy intakes demonstrated that the proportion of participants below the protein EAR was much lower (<5%) across all dietary groups, with the greatest change being for vegans. These findings may reflect the lower proportion of energy derived from protein in vegetarians and vegan diets, as well as the slightly higher rates of dietary under-reporting in these groups. This is supported by Mariotti *et al*. ^(28)^ who note under-reporting is an important methodological consideration when assessing the prevalence of inadequate protein intakes. Furthermore, the cut-off point used in the sensitivity analysis (EI:BMR ratio of 1.2) which reflects the minimum survival requirement, may still result in many under-reporters remaining in the data, and thus overestimate the prevalence of inadequate protein intake; applying a higher cut-off point more consistent with a sedentary lifestyle (EI:BMR ratio of 1.5) would likely further reduce the proportion of individuals falling below the EAR ^(18, 19)^.

Findings from FEED suggest that a large majority of vegans and vegetarians in the cohort have total protein intakes at or above the EAR, in line with previous literature^(4, 5, 23)^. With this said, the trend towards greater prevalence of inadequacy with more plant-based diets may be of concern when considering the potential for the lower protein quality of many plant-based proteins both in terms of digestibility (ie. how well the protein is broken down and absorbed in the gastrointestinal tract), and lower levels of some essential amino acids such as lysine and methionine ^(29)^. The EAR for protein is based principally on nitrogen balance and does not consider protein source digestibility ^(30)^.Thus, it is feasible that in those consuming the majority of their protein from homogenous lower quality plant sources such as grains ^(31)^, both the lower availability of amino acids for absorption in the digestive tract, and the relative inadequacies in certain essential amino acid may mean the EAR underestimates protein requirements. Compared to omnivores, the relative contributions of the top 10 contributing foods to total protein intake across other diets were lower (44.7% in omnivores vs 36.0%-37.3% in other diets), indicating the potential for a greater variety of protein sources in these diets. Thus, while vegans in FEED had a lower overall protein intake, they appear to consume a large variety of plant-based protein sources, not relying solely on grains, which may compensate for these factors. Additionally, many plant-based protein sources such as wholegrains, legumes and nuts are often accompanied by other healthful nutrients such as fiber, and so may be beneficial for public health outside of their protein content. The increased consumption of plant-based meat and dairy alternatives may also present new implications for public health; in the UK, analysis of survey data from online retailers in 2020 shows plant-based meat alternatives generally have a more healthful nutrient profile than meat equivalents, being lower in saturated fat, higher in fiber, but also contain additional salt ^(32)^. Additionally, the complete omission of animal foods may have implications for other nutrients of concern such as vitamin B12, although findings from FEED suggest that even non-supplementing vegans in the UK may not be deficient in vitamin B12, possibly due to increased availability of fortified products such as plant-based milks ^(33)^.

This study was strengthened by its large sample size for five dietary groups, and the inclusion of an updated FFQ for dietary data collection, including a number of contemporary plant-based protein alternatives. Nonetheless, certain limitations should be acknowledged. While FFQs are often used in large cohort studies to derive dietary data, they may lack precision for estimating absolute nutrient intakes which may result in under and overestimation of nutrient intakes ^(9, 11)^. Additionally, diet groups were self-reported and thus may be prone to some misclassification bias. Notably, a small amount of animal foods contribute to protein intake in vegan and vegetarian groups in the FEED study. We sought to minimise the spurious assignment of trace amounts of nutrients derived exclusively from animal-source foods to vegan participants by using plant-based alternatives in nutrient calculations where possible.

However, this could not be fully eliminated because of assumptions required when coding generic FFQ items and the potential for dietary reporting errors ^(5)^. The assignment of standard portion sizes to estimate nutritional intake may also result in some over/underestimation of serving sizes given these likely differ among individuals. The derivation of quantitative nutrient estimates from FFQs may also result in some inaccuracy in this study due to factors such as systematic error associated with such dietary assessment methods. As such this feature should be noted in the interpretation of these findings. Further, despite the use of the previously validated EPIC questionnaire, the modifications to include more questions on plant-based alternatives have not been validated currently. With this said the increased inclusion of plant-based alternatives and open text to allow for free reporting of foods in the questionnaire should increase the likelihood of capturing plant-based foods in the diet compared to previous studies. A large proportion of FEED participants are from the south of England, and are predominantly White women, with relatively high socioeconomic status ^(5)^. Thus, the data may not reflect accurately true population intakes across different regions of the UK, or be applicable across cultures.

## Conclusions

In this cohort of UK adults, protein sources differed substantially between diet groups. Grains were a major source of protein in all groups, as was dairy in all groups except vegans. Compared with omnivores, other diet groups obtained their protein from a wider range of foods, with no single food contributing as much to total intake. Plant-based foods were important sources of protein in these groups, particularly among vegetarians and vegans, where plant-based meat alternatives emerged as a leading source. Overall, seven percent of participants had protein intakes below the EAR, and this proportion progressively increased as more animal-source foods were excluded from the diet. However, this pattern was largely attenuated after excluding under-reporters. These findings highlight the emerging role of contemporary plant-based foods in supporting protein intake among people following plant-based diets. Future work should assess protein adequacy using biological measures, such as nitrogen balance), and by evaluating intakes of essential amino acids.

## Data Availability

The data access policy for the FEED study is available via the study website: https://www.ceu.ox.ac.uk/research/feeding-the-future-study-feed.

## Acknowledgments

We would like to thank the FEED participants for taking the time to complete the survey. We also wish to acknowledge the Nutrition Measurement Platform at the MRC Epidemiology Unit, University of Cambridge, for kindly sharing the UK National Diet and Nutrition Survey Nutrient Databank food composition database with us.

## Authorship

K.P. and I.L. contributed to conceptualization and methodology. I.L. conducted data curation and formal analysis, and contributed to visualization and writing (original draft preparation). W.B. contributed to data analysis, visualization and writing (original draft preparation). K.P. and T.K. contributed to supervision and writing (review & editing).

## Ethical Standards Disclosure

This study was conducted according to the guidelines laid down in the Declaration of Helsinki and all procedures involving research study participants were approved by the University of Oxford Medical Sciences Interdivisional Research Ethics Committee (MS IDREC). Electronic written informed consent was obtained from all subjects.

