## Supplementary Material for "Sources of protein in UK adults following contemporary plant-based diets: findings from the Feeding the Future (FEED) Study"

**Supplementary Table 1.** Food group definitions and percentage protein contribution to total protein intake by diet group

| **Source** | **food group** | **Individual survey items** | **Omnivore** | **Flexitarian** | **Pescatarian** | **Vegetarian** | **Vegan** |
| --- | --- | --- | --- | --- | --- | --- | --- |
| **Animal** | Dairy | cheese,  cheese sauce,  cream,  dairy full fat yogurt,  low-fat yogurt,  goat/sheep's milk,  other milk (dairy),  semi skimmed milk,  skimmed milk,  whole milk | 13.4 | 13.2 | 12.9 | 14.2 | 0.1 |
| **Animal** | Eggs | eggs | 5.7 | 4.5 | 5.4 | 5.4 | 0.2 |
| **Animal** | Fish | fish fried in batter/crumbs (e.g. fish burger, fish cakes, fish  fingers)  oily fish (e.g. salmon),  shellfish (e.g. prawns),  tuna,  other white fish | 11.2 | 12.5 | 14.9 | 0.3 | 0.1 |
| **Animal** | Meat | bacon,  unprocessed beef,  unprocessed lamb,  offal,  unprocessed pork,  unprocessed chicken, turkey,  other processed meat (e.g. nuggets, burgers, pie, sausage roll),  salami, dried meats (e.g. prosciutto),  sausages,  sandwich slices (e.g. pastrami, ham) | 33.0 | 13.4 | 0.3 | 0.1 | 0.0 |
| **Plant** | Fruit | apples, pears,  banana,  oranges, satsumas, grapefruit,  dried fruit (e.g. raisins, prunes (not in muesli)),  other fresh fruit (e.g. grapes, berries, kiwi),  tinned or stewed fruit | 1.6 | 2.5 | 2.7 | 2.8 | 3.5 |
| **Plant** | Grains | all-bran,  brown, granary, etc. bread or bread products,  brown rice,  couscous, bulgur wheat, etc,  crackers,  crispbread (e.g. Ryvita),  bran flakes, Weetabix, muesli,  oatcakes,  other bread products,  other breakfast cereal (e.g. cornflakes, Special K),  pizza,  porridge, ready brek, overnight oats,  sugary cereal (e.g. frosted flakes, crunchy granola),  white bread or bread products,  wholemeal bread or bread products,  white pasta/noodles,  wholemeal pasta,  white rice | 13.5 | 19.4 | 21.4 | 25.8 | 27.2 |
| **Plant** | Nuts/seeds | peanut/other nut butters (without sugar),  peanut/other nut butters (with added sugar),  nuts,  salted peanuts, other nuts or seeds,  tahini | 3.2 | 5.4 | 6.2 | 6.8 | 10.5 |
| **Plant** | Plant-based dairy alternatives | almond milk,  coconut milk,  hemp milk,  oat milk,  other milk alternatives,  pea milk,  rice milk,  soy milk,  vegan cheese,  vegan cream,  vegan yogurt | 0.4 | 1.3 | 2.0 | 3.0 | 7.0 |
| **Plant** | Plant-based meat alternatives | falafel,  Quorn, Quorn burgers  seitan,  soy burgers or sausages,  tempeh,  tofu, silken tofu,  textured vegetable protein (TVP, e.g. soy protein mince),  other veggie burgers or sausages | 1.3 | 4.6 | 8.3 | 12.0 | 15.0 |
| **Plant** | Potatoes/chips | chips, roast potatoes  boiled, mashed, or jacket potatoes | 1.5 | 1.6 | 1.9 | 2.3 | 2.3 |
| **Plant** | Pulses/legumes | baked beans,  chickpeas,  hummus,  other beans or lentils, | 2.2 | 4.5 | 6.0 | 6.8 | 10.5 |
| **Plant** | Vegetables | avocado,  cabbage, Brussels sprouts, cauliflower, coleslaw  carrot,  sweetcorn,  broccoli, spring greens, kale, spinach,  mushrooms,  onions, leeks  other vegetables (e.g. courgette, pumpkin),  peas, green beans,  sweet peppers,  parsnips, turnips, swedes, beetroot, other root vegetables,  salad vegetables (e.g. lettuce), cucumber,  tomatoes | 5.5 | 7.5 | 8.7 | 8.9 | 11.0 |
| **Mixed** | Other (discretionary) | beer, lager or cider,  butter,  chocolate spread,  cholesterol-lowering margarine,  crisps,  guacamole,  jam, marmalade, honey  lard/dripping,  low calorie soft drinks / diet fizzy soft drinks,  salad cream, dressing, dips, low-fat mayo (egg-based),  low-fat spread,  marmite,  mayonnaise,  meat soup,  olive oil,  other fat spread/margarine,  other sauces, white sauce, gravy (including vegan/vegetarian options),  other vegetable oil (e.g. rapeseed oil),  pure fruit juice or smoothie (excl. protein shake),  polyunsaturated margarine,  fizzy soft drinks e.g. coke,  spirits e.g. gin, whisky,  fruit squash or cordial,  sugar (added to tea, coffee, cereal),  tomato based sauce for pasta,  vegetable soup,  vegan salad cream, dressing, dips, low-fat mayo (non-egg-based),  vegan mayonnaise (non-egg-based, full-fat),  vegan fat spread/margarine (non-dairy),  water,  white hard vegetable fat (e.g. coconut),  wine, champagne, sherry, etc. | 3.3 | 3.7 | 3.7 | 4.6 | 4.5 |
| **Mixed** | Desserts and confectionery | biscuits (e.g. Digestive, chocolate-chip, Hobnob  cake),  cereal bar,  chocolate or sweets, all types,  fruit pie, fruit tart, crumble,  dairy ice cream, choc ices,  rice pudding, custard, creme caramel, mousse,  nut and/or dried fruit bar,  vegan ice cream, vegan choc ices | 2.5 | 3.2 | 3.2 | 4.1 | 2.8 |
| **Mixed** | Drinks containing milk/alternatives | hot chocolate, milkshake (incl. non-dairy),  coffee (all types),  milk-based smoothie,  tea (all types) | 0.8 | 1.0 | 1.1 | 1.2 | 1.0 |
| **Mixed** | Meal replacements | Huel shake/bar, other meal replacement shake/bar | 0.2 | 0.3 | 0.1 | 0.3 | 0.8 |
| **Mixed** | Protein shakes/bars | protein bar (hemp),  protein bar (other),  protein bar (pea),  protein bar (peanut),  protein bar (rice),  protein bar (soy),  protein bar (whey),  protein shake (hemp),  protein shake (other),  protein shake (pea),  protein shake (peanut),  protein shake (rice),  protein shake (soy),  protein shake (whey) | 1.0 | 1.4 | 1.5 | 1.6 | 3.4 |

### Supplementary Table 2. Mean daily intakes (g or ml) of all foods and beverages adjusted for age and gender and the respective protein contribution (%) by diet group in 6,113 FEED participants

| Food Item | Omnivore | | Flexitarian | | Pescatarian | | Vegetarian | | Vegan | |
| --- | --- | --- | --- | --- | --- | --- | --- | --- | --- | --- |
|  | Percentage of Total Protein (%) | Mean Intake (±SE) (g or ml) | Percentage of Total Protein (%) | Mean Intake (±SE) (g or ml) | Percentage of Total Protein (%) | Mean Intake (±SE) (g or ml) | Percentage of Total Protein (%) | Mean Intake (±SE) (g or ml) | Percentage of Total Protein (%) | Mean Intake (±SE) (g or ml) |
| All-Bran | 0.1 | 0.9 (±0.1) | 0.2 | 1.0 (±0.1) | 0.2 | 1.0 (±0.2) | 0.1 | 0.7 (±0.1) | 0.2 | 0.9 (±0.1) |
| Almond Milk | 0.0 | 7.3 (±1.4) | 0.1 | 10.1 (±1.5) | 0.1 | 14.5 (±2.4) | 0.1 | 11.3 (±1.6) | 0.1 | 13.6 (±1.5) |
| Apples, pears | 0.2 | 50.0 (±1.8) | 0.4 | 72.3 (±2.0) | 0.4 | 65.7 (±3.1) | 0.4 | 62.0 (±2.0) | 0.4 | 70.8 (±1.9) |
| Avocado | 0.4 | 22.9 (±0.9) | 0.6 | 26.9 (±1.0) | 0.8 | 32.4 (±1.6) | 0.6 | 23.4 (±1.1) | 0.8 | 28.9 (±1.0) |
| Bacon | 3.6 | 15.8 (±0.3) | 1.5 | 5.4 (±0.3) | 0.0 | 0.2 (±0.5) | 0.0 | 0.0 (±0.3) | 0.0 | 0.0 (±0.3) |
| Baked Beans | 0.6 | 12.5 (±0.5) | 0.7 | 13.6 (±0.6) | 1.0 | 17.6 (±0.9) | 1.3 | 19.5 (±0.6) | 1.7 | 23.7 (±0.5) |
| Banana | 0.4 | 31.3 (±1.1) | 0.6 | 43.8 (±1.2) | 0.7 | 45.2 (±1.9) | 0.7 | 42.1 (±1.3) | 0.9 | 54.5 (±1.2) |
| Beer, lager or cider | 0.1 | 45.8 (±2.8) | 0.1 | 48.2 (±3.1) | 0.1 | 56.8 (±4.8) | 0.2 | 55.0 (±3.2) | 0.2 | 46.3 (±2.9) |
| Biscuits | 0.3 | 4.8 (±0.2) | 0.4 | 5.5 (±0.3) | 0.4 | 5.2 (±0.4) | 0.5 | 6.2 (±0.3) | 0.4 | 4.6 (±0.2) |
| Boiled, mashed, or jacket potatoes | 0.9 | 44.7 (±1.1) | 1.0 | 44.3 (±1.2) | 1.1 | 43.1 (±1.9) | 1.4 | 49.4 (±1.3) | 1.3 | 46.0 (±1.1) |
| Bran flakes, Weetabix, muesli | 0.9 | 8.3 (±0.4) | 1.4 | 11.2 (±0.5) | 1.5 | 10.5 (±0.7) | 1.6 | 10.3 (±0.5) | 1.4 | 9.2 (±0.4) |
| Broccoli, spring greens, kale, spinach | 1.0 | 40.5 (±1) | 1.4 | 44.9 (±1.1) | 1.6 | 49.0 (±1.7) | 1.5 | 41.2 (±1.1) | 2.2 | 57.2 (±1.0) |
| Brown rice | 0.4 | 11.9 (±0.6) | 0.8 | 19.4 (±0.7) | 0.9 | 20.5 (±1.1) | 1.0 | 19.3 (±0.7) | 1.4 | 27.6 (±0.6) |
| Brown/granary bread or bread products | 1.9 | 18.4 (±0.9) | 2.7 | 23.3 (±1.0) | 3.0 | 23.4 (±1.5) | 3.5 | 24.0 (±1.0) | 3.4 | 23.8 (±0.9) |
| Butter | 0.0 | 7.8 (±0.2) | 0.0 | 4.8 (±0.2) | 0.0 | 3.9 (±0.3) | 0.0 | 3.6 (±0.2) | 0.0 | 0.3 (±0.2) |
| Cabbage, Brussels sprouts, Cauliflower, coleslaw | 0.5 | 26.1 (±0.7) | 0.6 | 26.8 (±0.8) | 0.7 | 27.6 (±1.2) | 0.8 | 25.5 (±0.8) | 1.0 | 34.1 (±0.7) |
| Cake | 0.8 | 14.7 (±0.5) | 1.1 | 16.5 (±0.5) | 1.0 | 13.1 (±0.8) | 1.3 | 15.7 (±0.5) | 0.7 | 10.9 (±0.5) |
| Carrot | 0.1 | 22.0 (±0.5) | 0.2 | 26.6 (±0.6) | 0.2 | 25.1 (±0.9) | 0.2 | 24.8 (±0.6) | 0.2 | 30.2 (±0.6) |
| Cereal Bar | 0.1 | 2.2 (±0.1) | 0.2 | 2.4 (±0.1) | 0.2 | 2.5 (±0.2) | 0.2 | 2.8 (±0.1) | 0.2 | 2.1 (±0.1) |
| Cheese | 3.7 | 20.9 (±0.4) | 3.7 | 17.0 (±0.4) | 3.9 | 16.8 (±0.7) | 5.2 | 19.7 (±0.5) | 0.0 | 0.0 (±0.4) |
| Cheese Sauce | 0.5 | 5.0 (±0.1) | 0.5 | 3.9 (±0.1) | 0.5 | 4.3 (±0.2) | 0.6 | 4.2 (±0.2) | 0.0 | 0.2 (±0.1) |
| Chickpeas | 0.5 | 7.9 (±0.5) | 1.3 | 15.5 (±0.6) | 1.8 | 20.3 (±0.9) | 1.9 | 18.5 (±0.6) | 3.0 | 28.7 (±0.5) |
| Chips, roast potatoes | 0.6 | 25.9 (±0.6) | 0.6 | 21.2 (±0.7) | 0.7 | 25.3 (±1.1) | 0.9 | 26.7 (±0.7) | 1.0 | 28.2 (±0.7) |
| Chocolate Spread | 0.0 | 0.8 (±0.1) | 0.0 | 0.8 (±0.1) | 0.0 | 0.8 (±0.1) | 0.1 | 0.9 (±0.1) | 0.0 | 0.6 (±0.1) |
| Chocolate or sweets, all types | 0.3 | 9.6 (±0.4) | 0.4 | 10.6 (±0.4) | 0.4 | 11.0 (±0.6) | 0.6 | 12.1 (±0.4) | 0.4 | 8.3 (±0.4) |
| Cholesterol-lowering margarine | 0.0 | 0.2 (±0.0) | 0.0 | 0.2 (±0.0) | 0.0 | 0.4 (±0.1) | 0.0 | 0.4 (±0) | 0.0 | 0.1 (±0.0) |
| Coconut Milk | 0.0 | 2.2 (±0.8) | 0.0 | 2.8 (±0.9) | 0.0 | 7.3 (±1.4) | 0.0 | 3.3 (±0.9) | 0.0 | 5.2 (±0.8) |
| Coffee, all types | 0.4 | 267.9 (±5.8) | 0.5 | 267.0 (±6.3) | 0.5 | 253.6 (±9.8) | 0.6 | 245.8 (±6.6) | 0.6 | 250.8 (±6.0) |
| Couscous, bulgur wheat, etc | 0.4 | 8.6 (±0.5) | 0.8 | 15.0 (±0.5) | 1.0 | 17.7 (±0.8) | 0.9 | 14.6 (±0.5) | 1.2 | 17.7 (±0.5) |
| Crackers | 0.2 | 1.9 (±0.1) | 0.2 | 2.1 (±0.1) | 0.3 | 2.2 (±0.2) | 0.3 | 2.3 (±0.1) | 0.2 | 1.9 (±0.1) |
| Cream | 0.2 | 7.0 (±0.2) | 0.1 | 2.0 (±0.2) | 0.1 | 1.8 (±0.4) | 0.1 | 1.6 (±0.3) | 0.0 | 0.1 (±0.2) |
| Crispbread | 0.1 | 1.2 (±0.1) | 0.2 | 1.9 (±0.1) | 0.2 | 1.8 (±0.2) | 0.2 | 1.6 (±0.1) | 0.2 | 1.5 (±0.1) |
| Crisps | 0.4 | 7.9 (±0.3) | 0.4 | 7.5 (±0.3) | 0.5 | 8.3 (±0.5) | 0.7 | 10.1 (±0.3) | 0.7 | 8.7 (±0.3) |
| Dairy full fat yogurt | 1.4 | 29.9 (±0.9) | 1.7 | 29.2 (±1.0) | 1.4 | 21.6 (±1.5) | 1.5 | 20.6 (±1.0) | 0.0 | 1.2 (±0.9) |
| Dairy ice cream | 0.3 | 11.6 (±0.3) | 0.3 | 9.9 (±0.4) | 0.3 | 8.2 (±0.5) | 0.4 | 9.2 (±0.4) | 0.0 | 0.5 (±0.3) |
| Dried fruit | 0.2 | 10.0 (±0.6) | 0.4 | 15.5 (±0.6) | 0.5 | 16.4 (±1.0) | 0.5 | 13.2 (±0.7) | 0.7 | 20.0 (±0.6) |
| Dried meats | 0.9 | 3.7 (±0.1) | 0.5 | 1.9 (±0.1) | 0.0 | 0.1 (±0.1) | 0.0 | 0.0 (±0.1) | 0.0 | 0.0 (±0.1) |
| Eggs | 5.7 | 47.3 (±1.1) | 4.5 | 32.6 (±1.3) | 5.4 | 37.5 (±2) | 5.4 | 34.1 (±1.4) | 0.2 | 1.0 (±1.1) |
| Falafel | 0.3 | 3.5 (±0.2) | 0.6 | 6.9 (±0.2) | 0.8 | 8.4 (±0.4) | 0.9 | 8.4 (±0.2) | 1.1 | 8.9 (±0.2) |
| Fish fried in batter/crumbs | 1.0 | 7.9 (±0.1) | 1.2 | 7.5 (±0.2) | 1.6 | 9.5 (±0.2) | 0.0 | 0.3 (±0.2) | 0.0 | 0.1 (±0.2) |
| Fizzy soft drinks | 0.0 | 20.5 (±1.5) | 0.0 | 12.1 (±1.6) | 0.0 | 14.9 (±2.5) | 0.0 | 15.8 (±1.7) | 0.0 | 12.5 (±1.5) |
| Fruit pie, fruit tart, crumble | 0.1 | 3.9 (±0.1) | 0.1 | 4.0 (±0.1) | 0.1 | 3.9 (±0.2) | 0.2 | 4.4 (±0.1) | 0.1 | 3.7 (±0.1) |
| Fruit squash or cordial | 0.0 | 108.3 (±4.9) | 0.0 | 66 (±5.3) | 0.0 | 72.9 (±8.3) | 0.0 | 83.9 (±5.6) | 0.0 | 78.1 (±5.1) |
| Goat/sheep's milk | 0.1 | 1.7 (±0.4) | 0.1 | 1.4 (±0.4) | 0.1 | 1.3 (±0.7) | 0.0 | 0.8 (±0.4) | 0.0 | 0.1 (±0.4) |
| Guacamole | 0.0 | 1.6 (±0.1) | 0.0 | 1.6 (±0.1) | 0.0 | 1.9 (±0.2) | 0.0 | 2.0 (±0.1) | 0.1 | 2.1 (±0.1) |
| Hemp Milk | 0.0 | 0.0 (±0.2) | 0.0 | 0.2 (±0.2) | 0.0 | 0.3 (±0.4) | 0.0 | 0.3 (±0.3) | 0.0 | 0.9 (±0.2) |
| Hot chocolate, milkshake | 0.1 | 15.6 (±1.0) | 0.1 | 15.0 (±1.0) | 0.2 | 16.6 (±1.6) | 0.2 | 19.2 (±1.1) | 0.2 | 15.5 (±1.0) |
| Huel shake/bar | 0.1 | 5.2 (±2.7) | 0.1 | 4.4 (±2.6) | 0.1 | 3.7 (±4.1) | 0.2 | 10.4 (±2.8) | 0.6 | 14.3 (±1.9) |
| Hummus | 0.3 | 3.6 (±0.3) | 0.6 | 6.6 (±0.3) | 0.7 | 7.9 (±0.5) | 0.9 | 8.2 (±0.4) | 1.5 | 13.8 (±0.3) |
| Jam, marmalade, honey | 0.0 | 4.4 (±0.2) | 0.0 | 5.4 (±0.2) | 0.0 | 5.1 (±0.3) | 0.0 | 4.9 (±0.2) | 0.0 | 3.9 (±0.2) |
| Lard/dripping | 0.0 | 1.5 (±0.1) | 0.0 | 0.1 (±0.1) | 0.0 | 0.0 (±0.1) | 0.0 | 0.0 (±0.1) | 0.0 | 0.0 (±0.1) |
| Low calorie soft drinks | 0.0 | 73.0 (±4.3) | 0.0 | 48.0 (±4.6) | 0.0 | 51.8 (±7.2) | 0.0 | 73.1 (±4.8) | 0.0 | 49.8 (±4.4) |
| Low-fat spread | 0.0 | 0.4 (±0.0) | 0.0 | 0.4 (±0.0) | 0.0 | 0.3 (±0.1) | 0.0 | 0.3 (±0.0) | 0.0 | 0.1 (±0.0) |
| Low-fat yogurt | 1.1 | 19.7 (±1.0) | 1.7 | 26.4 (±1.1) | 1.9 | 27.7 (±1.7) | 1.7 | 21.3 (±1.1) | 0.0 | 0.6 (±1.0) |
| Marmite | 0.3 | 0.9 (±0.1) | 0.5 | 1.2 (±0.1) | 0.6 | 1.4 (±0.1) | 0.9 | 1.9 (±0.1) | 1.2 | 2.5 (±0.1) |
| Mayonnaise | 0.1 | 5.9 (±0.2) | 0.1 | 4.2 (±0.2) | 0.1 | 4.2 (±0.3) | 0.0 | 3.0 (±0.2) | 0.0 | 0.1 (±0.2) |
| Meat Soup | 1.0 | 16.3 (±0.4) | 0.7 | 8.9 (±0.4) | 0.3 | 3.3 (±0.7) | 0.0 | 0.1 (±0.4) | 0.0 | 0.0 (±0.4) |
| Meat alternatives | 0.1 | 0.5 (±0.3) | 0.2 | 1.2 (±0.3) | 0.7 | 2.9 (±0.5) | 1.1 | 4.1 (±0.3) | 1.6 | 6.0 (±0.3) |
| Milk-based pudding | 0.3 | 9.1 (±0.3) | 0.3 | 8.0 (±0.4) | 0.3 | 7.5 (±0.6) | 0.4 | 8.5 (±0.4) | 0.2 | 4.5 (±0.4) |
| Milk-based smoothie | 0.2 | 8.1 (±0.9) | 0.3 | 8.9 (±0.9) | 0.3 | 9.3 (±1.5) | 0.3 | 8.1 (±1.0) | 0.1 | 13.0 (±0.9) |
| Mushrooms | 0.2 | 12.7 (±0.4) | 0.2 | 13.8 (±0.4) | 0.3 | 15.7 (±0.7) | 0.3 | 15.7 (±0.4) | 0.4 | 20.4 (±0.4) |
| Nut and/or dried fruit bar | 0.2 | 2.3 (±0.2) | 0.3 | 3.1 (±0.2) | 0.4 | 3.7 (±0.3) | 0.4 | 3.4 (±0.2) | 0.5 | 4.3 (±0.2) |
| Nut butters (with added sugar) | 0.1 | 0.5 (±0.1) | 0.1 | 0.5 (±0.1) | 0.2 | 0.7 (±0.1) | 0.3 | 0.9 (±0.1) | 0.4 | 1.2 (±0.1) |
| Nut butters (without sugar) | 0.6 | 2.9 (±0.3) | 1.2 | 4.9 (±0.3) | 1.4 | 5.6 (±0.4) | 1.5 | 5 (±0.3) | 2.8 | 8.9 (±0.3) |
| Nuts | 1.7 | 8.3 (±0.5) | 2.9 | 12.0 (±0.5) | 3.1 | 11.9 (±0.8) | 3.4 | 11.3 (±0.5) | 5.0 | 17.4 (±0.5) |
| Oat Milk | 0.1 | 16.4 (±2.9) | 0.4 | 49.4 (±3.1) | 0.5 | 62.5 (±4.9) | 0.6 | 65 (±3.3) | 1.0 | 110.6 (±3.0) |
| Oatcakes | 0.2 | 1.8 (±0.1) | 0.4 | 2.9 (±0.2) | 0.4 | 2.9 (±0.2) | 0.3 | 2.2 (±0.2) | 0.3 | 2.5 (±0.1) |
| Offal | 0.9 | 4.5 (±0.2) | 0.3 | 1.3 (±0.2) | 0.0 | 0.1 (±0.3) | 0.0 | 0.0 (±0.2) | 0.0 | 0.0 (±0.2) |
| Oily fish | 3.7 | 16.9 (±0.4) | 4.3 | 16.7 (±0.4) | 5.1 | 18.2 (±0.6) | 0.1 | 0.2 (±0.4) | 0.0 | 0.4 (±0.4) |
| Olive Oil | 0.0 | 5.8 (±0.2) | 0.0 | 7.5 (±0.2) | 0.0 | 6.6 (±0.3) | 0.0 | 5.9 (±0.2) | 0.0 | 5.7 (±0.2) |
| Onions, leeks | 0.6 | 37.2 (±0.8) | 0.8 | 41.2 (±0.9) | 0.9 | 42.8 (±1.4) | 1.1 | 42.4 (±0.9) | 1.2 | 49.1 (±0.9) |
| Oranges, satsumas, grapefruit | 0.3 | 35.3 (±1.4) | 0.4 | 46.3 (±1.5) | 0.4 | 42.7 (±2.4) | 0.4 | 40.1 (±1.6) | 0.5 | 49.3 (±1.5) |
| Other Milk (Dairy) | 0.2 | 6.3 (±0.8) | 0.2 | 5.2 (±0.9) | 0.0 | 1 (±1.4) | 0.1 | 2.2 (±0.9) | 0.0 | 0.2 (±0.8) |
| Other beans or lentils | 0.8 | 11.7 (±0.7) | 1.9 | 23.7 (±0.8) | 2.4 | 28.2 (±1.2) | 2.7 | 27.5 (±0.8) | 4.4 | 43.3 (±0.7) |
| Other bread products | 0.4 | 5.5 (±0.2) | 0.5 | 5.9 (±0.2) | 0.6 | 6.2 (±0.4) | 0.7 | 7.0 (±0.3) | 0.7 | 6.4 (±0.2) |
| Other breakfast cereal | 0.3 | 3.2 (±0.2) | 0.3 | 2.8 (±0.2) | 0.2 | 2.0 (±0.3) | 0.4 | 3.2 (±0.2) | 0.3 | 2.6 (±0.2) |
| Other fat spread/margarine | 0.0 | 0.4 (±0.0) | 0.0 | 0.5 (±0.1) | 0.0 | 0.3 (±0.1) | 0.0 | 0.3 (±0.1) | 0.0 | 0.1 (±0.1) |
| Other fresh fruit | 0.4 | 48.0 (±1.7) | 0.7 | 62.3 (±1.8) | 0.7 | 59.3 (±2.8) | 0.7 | 54.0 (±1.9) | 0.8 | 65.4 (±1.7) |
| Other meal replacement shake/bar | 0.1 | 9.9 (±2.5) | 0.2 | 12.7 (±2.4) | 0.1 | 3.4 (±3.7) | 0.1 | 4.5 (±2.6) | 0.2 | 1.4 (±1.7) |
| Other milk alternatives | 0.0 | 1.4 (±0.7) | 0.0 | 2.3 (±0.8) | 0.0 | 3.0 (±1.2) | 0.0 | 2.4 (±0.8) | 0.0 | 3.4 (±0.7) |
| Other processed meat | 1.0 | 7.3 (±0.1) | 0.6 | 3.6 (±0.2) | 0.0 | 0.3 (±0.2) | 0.0 | 0.1 (±0.2) | 0.0 | 0.0 (±0.1) |
| Other sauces, white sauce, gravy | 0.1 | 6.7 (±0.2) | 0.1 | 4.9 (±0.2) | 0.1 | 4.8 (±0.3) | 0.2 | 6.3 (±0.2) | 0.0 | 4.6 (±0.2) |
| Other vegetable oil | 0.0 | 2.3 (±0.1) | 0.0 | 2.7 (±0.1) | 0.0 | 2.6 (±0.2) | 0.0 | 3.0 (±0.1) | 0.0 | 3.3 (±0.1) |
| Other vegetables | 0.3 | 28.2 (±1.0) | 0.5 | 36.0 (±1.0) | 0.5 | 34.3 (±1.6) | 0.6 | 33.8 (±1.1) | 0.8 | 42.5 (±1) |
| Other veggie burgers or sausages | 0.2 | 1.9 (±0.2) | 0.9 | 5.8 (±0.2) | 1.5 | 8.9 (±0.4) | 2.1 | 11.1 (±0.3) | 2.1 | 10.3 (±0.2) |
| Pea Milk | 0.0 | 0.1 (±0.2) | 0.0 | 0.2 (±0.2) | 0.0 | 0.0 (±0.3) | 0.0 | 0.0 (±0.2) | 0.0 | 0.3 (±0.2) |
| Peas, green beans | 1.1 | 23.5 (±0.6) | 1.5 | 26.6 (±0.6) | 1.8 | 28.7 (±1.0) | 1.8 | 25.4 (±0.7) | 2.2 | 30.8 (±0.6) |
| Pizza | 1.3 | 11.8 (±0.4) | 1.7 | 12.4 (±0.5) | 1.9 | 13.6 (±0.8) | 2.5 | 15.3 (±0.5) | 2.9 | 17.8 (±0.5) |
| Polyunsaturated margarine | 0.0 | 0.3 (±0.0) | 0.0 | 0.3 (±0.0) | 0.0 | 0.4 (±0.1) | 0.0 | 0.4 (±0.0) | 0.0 | 0.2 (±0.0) |
| Porridge, Ready brek, Overnight Oats | 0.6 | 41.2 (±1.9) | 1.0 | 62.1 (±2.0) | 1.2 | 70.5 (±3.2) | 1.1 | 57.3 (±2.1) | 1.4 | 73 (±1.9) |
| Protein Bar (Hemp) | 0.0 | 0.0 (±0.0) | 0.0 | 0.0 (±0.0) | 0.0 | 0.0 (±0.0) | 0.0 | 0.0 (±0.0) | 0.0 | 0.0 (±0.0) |
| Protein Bar (Other) | 0.0 | 0.2 (±0.1) | 0.0 | 0.1 (±0.1) | 0.1 | 0.2 (±0.1) | 0.1 | 0.2 (±0.1) | 0.2 | 0.5 (±0.1) |
| Protein Bar (Pea) | 0.0 | 0.1 (±0.1) | 0.1 | 0.3 (±0.1) | 0.1 | 0.2 (±0.1) | 0.1 | 0.2 (±0.1) | 0.2 | 0.4 (±0.1) |
| Protein Bar (Peanut) | 0.0 | 0.0 (±0.0) | 0.0 | 0.0 (±0.0) | 0.0 | 0.1 (±0.0) | 0.0 | 0.1 (±0.0) | 0.0 | 0.0 (±0.0) |
| Protein Bar (Rice) | 0.0 | 0.0 (±0.0) | 0.0 | 0.0 (±0.0) | 0.0 | 0.0 (±0.0) | 0.0 | 0.0 (±0.0) | 0.0 | 0.0 (±0.0) |
| Protein Bar (Soy) | 0.0 | 0.0 (±0.1) | 0.0 | 0.1 (±0.1) | 0.1 | 0.2 (±0.1) | 0.1 | 0.2 (±0.1) | 0.2 | 0.4 (±0.1) |
| Protein Bar (Whey) | 0.1 | 0.5 (±0.1) | 0.2 | 0.5 (±0.1) | 0.1 | 0.2 (±0.1) | 0.1 | 0.2 (±0.1) | 0.0 | 0.0 (±0.1) |
| Protein Shake (Hemp) | 0.0 | 0.0 (±0.0) | 0.0 | 0.0 (±0.0) | 0.0 | 0.1 (±0.1) | 0.0 | 0.0 (±0.0) | 0.1 | 0.1 (±0.0) |
| Protein Shake (Other) | 0.1 | 0.2 (±0.1) | 0.1 | 0.2 (±0.1) | 0.2 | 0.2 (±0.1) | 0.3 | 0.3 (±0.1) | 0.6 | 0.6 (±0.1) |
| Protein Shake (Pea) | 0.1 | 0.1 (±0.1) | 0.3 | 0.3 (±0.1) | 0.4 | 0.4 (±0.2) | 0.4 | 0.4 (±0.1) | 1.4 | 1.1 (±0.1) |
| Protein Shake (Peanut) | 0.0 | 0.0 (±0.0) | 0.0 | 0.0 (±0.0) | 0.0 | 0.0 (±0.0) | 0.0 | 0.0 (±0.0) | 0.0 | 0.0 (±0.0) |
| Protein Shake (Rice) | 0.0 | 0.0 (±0.0) | 0.0 | 0.0 (±0.0) | 0.0 | 0.0 (±0.0) | 0.0 | 0.0 (±0.0) | 0.0 | 0.0 (±0.0) |
| Protein Shake (Soy) | 0.0 | 0.0 (±0.1) | 0.2 | 0.2 (±0.1) | 0.2 | 0.2 (±0.1) | 0.2 | 0.2 (±0.1) | 0.7 | 0.5 (±0.1) |
| Protein Shake (Whey) | 0.6 | 0.8 (±0.1) | 0.5 | 0.5 (±0.1) | 0.4 | 0.5 (±0.2) | 0.4 | 0.4 (±0.1) | 0.0 | 0.0 (±0.1) |
| Pure fruit juice or smoothie | 0.2 | 49.0 (±2.5) | 0.3 | 48.8 (±2.7) | 0.3 | 45.1 (±4.3) | 0.4 | 58.7 (±2.9) | 0.4 | 54 (±2.6) |
| Quorn | 0.2 | 1.9 (±0.3) | 0.8 | 5.8 (±0.4) | 1.9 | 12.3 (±0.6) | 3.0 | 16.8 (±0.4) | 1.3 | 6.9 (±0.3) |
| Rice Milk | 0.0 | 0.3 (±0.3) | 0.0 | 0.9 (±0.3) | 0.0 | 0.1 (±0.5) | 0.0 | 1.1 (±0.4) | 0.0 | 0.7 (±0.3) |
| Root vegetables | 0.2 | 14.2 (±0.5) | 0.2 | 17.4 (±0.5) | 0.2 | 17.2 (±0.8) | 0.3 | 16.4 (±0.5) | 0.3 | 20.4 (±0.5) |
| Salad cream, dressing, dips, low-fat mayo | 0.0 | 2.1 (±0.1) | 0.0 | 1.8 (±0.1) | 0.0 | 1.7 (±0.2) | 0.0 | 1.7 (±0.1) | 0.0 | 0.1 (±0.1) |
| Salad vegetables | 0.3 | 31.5 (±0.9) | 0.5 | 37.8 (±0.9) | 0.5 | 37.1 (±1.5) | 0.5 | 33.4 (±1) | 0.6 | 40.0 (±0.9) |
| Salted peanuts, other nuts or seeds | 0.6 | 3.2 (±0.2) | 0.8 | 3.7 (±0.2) | 0.9 | 4.0 (±0.3) | 1.0 | 3.9 (±0.2) | 1.2 | 4.6 (±0.2) |
| Sandwich slices | 0.7 | 3.3 (±0.1) | 0.3 | 1.2 (±0.1) | 0.0 | 0.0 (±0.1) | 0.0 | 0.0 (±0.1) | 0.0 | 0.0 (±0.1) |
| Sausages | 1.0 | 6.9 (±0.1) | 0.6 | 3.3 (±0.1) | 0.0 | 0.1 (±0.2) | 0.0 | 0.0 (±0.1) | 0.0 | 0.0 (±0.1) |
| Seitan | 0.0 | 0.2 (±0.2) | 0.2 | 0.8 (±0.2) | 0.3 | 1.5 (±0.3) | 0.5 | 2.0 (±0.2) | 1.5 | 5.3 (±0.2) |
| Semi Skimmed Milk | 3.6 | 106.6 (±2.9) | 3.6 | 88.1 (±3.2) | 3.2 | 71.5 (±4.9) | 3.0 | 59.6 (±3.3) | 0.0 | 2.8 (±3.0) |
| Shellfish | 0.9 | 4.8 (±0.1) | 0.9 | 3.9 (±0.1) | 1.1 | 4.5 (±0.2) | 0.0 | 0.1 (±0.1) | 0.0 | 0.0 (±0.1) |
| Skimmed Milk | 0.8 | 23.0 (±1.9) | 1.1 | 26.1 (±2.1) | 1.2 | 26.2 (±3.2) | 1.4 | 27.2 (±2.2) | 0.0 | 1.6 (±2.0) |
| Soy Milk | 0.2 | 6.8 (±2.7) | 0.6 | 17.8 (±2.9) | 1.0 | 26.5 (±4.6) | 1.7 | 38.6 (±3.1) | 4.3 | 97.1 (±2.8) |
| Soya burgers or sausages | 0.1 | 0.7 (±0.2) | 0.4 | 2.3 (±0.2) | 0.7 | 3.9 (±0.3) | 1.1 | 5.3 (±0.2) | 1.5 | 6.8 (±0.2) |
| Spirits | 0.0 | 3.5 (±0.2) | 0.0 | 3.0 (±0.2) | 0.0 | 3.2 (±0.3) | 0.0 | 3.2 (±0.2) | 0.0 | 2.6 (±0.2) |
| Sugar | 0.0 | 1.2 (±0.1) | 0.0 | 0.8 (±0.1) | 0.0 | 0.6 (±0.1) | 0.0 | 0.9 (±0.1) | 0.0 | 1.0 (±0.1) |
| Sugary cereal | 0.2 | 2.7 (±0.2) | 0.2 | 2.7 (±0.2) | 0.2 | 2.5 (±0.3) | 0.3 | 3.2 (±0.2) | 0.2 | 1.8 (±0.2) |
| Sweet peppers | 0.3 | 32.0 (±0.9) | 0.4 | 37.3 (±1.0) | 0.5 | 43.3 (±1.6) | 0.5 | 38.1 (±1.1) | 0.5 | 42.3 (±1.0) |
| Sweetcorn | 0.1 | 3.2 (±0.1) | 0.1 | 3.5 (±0.1) | 0.1 | 3.8 (±0.2) | 0.1 | 3.9 (±0.1) | 0.2 | 4.5 (±0.1) |
| Tahini | 0.2 | 1.2 (±0.1) | 0.5 | 2.1 (±0.2) | 0.5 | 2.3 (±0.2) | 0.7 | 2.6 (±0.2) | 1.1 | 4.2 (±0.2) |
| Tea, all types | 0.1 | 341.0 (±6.7) | 0.1 | 357.5 (±7.3) | 0.1 | 364.5 (±11.3) | 0.1 | 343.6 (±7.6) | 0.1 | 338.0 (±6.9) |
| Tempeh | 0.1 | 0.4 (±0.2) | 0.4 | 1.5 (±0.2) | 0.6 | 2.5 (±0.3) | 0.8 | 2.6 (±0.2) | 1.9 | 6.2 (±0.2) |
| Textured vegetable protein | 0.1 | 0.2 (±0.0) | 0.4 | 0.7 (±0.1) | 0.7 | 1.3 (±0.1) | 1.1 | 1.6 (±0.1) | 1.2 | 1.7 (±0.0) |
| Tinned or stewed fruit | 0.0 | 9.1 (±0.6) | 0.1 | 9.8 (±0.6) | 0.1 | 8.4 (±0.9) | 0.1 | 9.2 (±0.6) | 0.1 | 8.4 (±0.6) |
| Tofu | 0.2 | 2.3 (±0.4) | 0.7 | 7.3 (±0.4) | 1.0 | 10.4 (±0.7) | 1.3 | 11.4 (±0.5) | 2.8 | 23.4 (±0.4) |
| Tomato based sauce | 0.2 | 13.3 (±0.4) | 0.3 | 14.7 (±0.4) | 0.3 | 15.1 (±0.7) | 0.4 | 16.8 (±0.4) | 0.4 | 16.8 (±0.4) |
| Tomatoes | 0.4 | 47.8 (±1.1) | 0.5 | 56.0 (±1.2) | 0.6 | 53.6 (±1.9) | 0.7 | 56.4 (±1.3) | 0.7 | 60.0 (±1.1) |
| Tuna | 2.7 | 9.7 (±0.2) | 3.0 | 9.0 (±0.2) | 3.5 | 9.8 (±0.3) | 0.1 | 0.2 (±0.2) | 0.0 | 0.0 (±0.2) |
| Unprocessed beef | 8.6 | 33.4 (±0.7) | 2.2 | 7.2 (±0.8) | 0.0 | 0.4 (±1.2) | 0.0 | 0.1 (±0.8) | 0.0 | 0.0 (±0.8) |
| Unprocessed chicken, turkey | 8.7 | 32.4 (±0.4) | 4.6 | 14.4 (±0.4) | 0.2 | 0.6 (±0.7) | 0.0 | 0.1 (±0.5) | 0.0 | 0.0 (±0.4) |
| Unprocessed lamb | 3.4 | 13.3 (±0.3) | 1.4 | 4.5 (±0.3) | 0.0 | 0.0 (±0.5) | 0.0 | 0.0 (±0.3) | 0.0 | 0.1 (±0.3) |
| Unprocessed pork | 4.4 | 17.0 (±0.3) | 1.4 | 4.8 (±0.4) | 0.0 | 0.0 (±0.6) | 0.0 | 0.0 (±0.4) | 0.0 | 0.0 (±0.3) |
| Vegan cheese | 0.0 | 0.4 (±0.1) | 0.0 | 1.0 (±0.2) | 0.1 | 1.7 (±0.2) | 0.1 | 2.6 (±0.2) | 0.4 | 8.3 (±0.2) |
| Vegan cream | 0.0 | 0.2 (±0.1) | 0.0 | 0.6 (±0.1) | 0.0 | 0.7 (±0.1) | 0.0 | 1.0 (±0.1) | 0.0 | 2.1 (±0.1) |
| Vegan fat spread/margarine | 0.1 | 2.5 (±0.8) | 0.2 | 7.4 (±0.9) | 0.3 | 9.3 (±1.3) | 0.4 | 13.6 (±0.9) | 1.2 | 36.1 (±0.8) |
| Vegan ice cream | 0.0 | 1.7 (±0.3) | 0.0 | 3.1 (±0.3) | 0.1 | 5.0 (±0.5) | 0.1 | 5.8 (±0.4) | 0.2 | 12.6 (±0.3) |
| Vegan mayonnaise | 0.0 | 0.4 (±0.1) | 0.0 | 0.9 (±0.1) | 0.0 | 1.5 (±0.2) | 0.0 | 2.3 (±0.1) | 0.0 | 5.0 (±0.1) |
| Vegan salad cream, dressing, dips, low-fat mayo | 0.0 | 0.1 (±0.1) | 0.0 | 0.2 (±0.1) | 0.0 | 0.5 (±0.1) | 0.0 | 0.9 (±0.1) | 0.0 | 1.8 (±0.1) |
| Vegan salad cream, dressing, dips, low-fat mayo | 0.0 | 0.2 (±0.1) | 0.0 | 0.8 (±0.1) | 0.0 | 1.1 (±0.2) | 0.0 | 2.6 (±0.1) | 0.0 | 5.4 (±0.1) |
| Vegan yogurt | 0.0 | 643.9 (±8.4) | 0.0 | 696.2 (±9.1) | 0.0 | 666.8 (±14.1) | 0.0 | 627.7 (±9.5) | 0.0 | 661.3 (±8.6) |
| Vegetable soup | 0.6 | 29.5 (±1.2) | 1.0 | 42.3 (±1.3) | 1.1 | 41.2 (±2.0) | 1.4 | 45.5 (±1.3) | 1.4 | 46.7 (±1.2) |
| Water | 1.4 | 17.2 (±0.7) | 1.2 | 12.4 (±0.8) | 1.2 | 11.2 (±1.3) | 1.9 | 15.9 (±0.8) | 1.7 | 13.0 (±0.8) |
| White bread or bread products | 0.0 | 0.6 (±0.0) | 0.0 | 0.3 (±0.0) | 0.0 | 0.4 (±0.1) | 0.0 | 0.3 (±0.0) | 0.0 | 0.5 (±0) |
| White hard vegetable fat | 2.3 | 23.6 (±1.1) | 3.5 | 32.6 (±1.2) | 4.0 | 33.0 (±1.9) | 5.1 | 36.8 (±1.3) | 5.4 | 40.9 (±1.2) |
| White pasta/noodles | 1.6 | 33.5 (±1.0) | 2.1 | 36.9 (±1.1) | 2.2 | 37.1 (±1.7) | 2.8 | 40.1 (±1.1) | 2.5 | 32.4 (±1.0) |
| White rice | 0.7 | 25.2 (±0.7) | 0.8 | 24.3 (±0.8) | 0.8 | 23.4 (±1.2) | 0.9 | 24.0 (±0.8) | 1.0 | 22.8 (±0.7) |
| Whole Milk | 1.9 | 57.2 (±2.2) | 0.8 | 21.6 (±2.3) | 0.7 | 17.1 (±3.6) | 0.6 | 13.0 (±2.4) | 0.0 | 0.0 (±2.2) |
| Wholemeal bread or bread products | 2.9 | 12.7 (±0.3) | 3.2 | 11.6 (±0.3) | 3.7 | 12.4 (±0.4) | 0.1 | 0.2 (±0.3) | 0.0 | 0.3 (±0.3) |
| Wholemeal pasta | 0.7 | 14.8 (±0.8) | 1.5 | 24.8 (±0.9) | 1.7 | 27.5 (±1.4) | 1.9 | 26.7 (±1.0) | 2.6 | 35.3 (±0.9) |
| Wine, champagne, sherry | 0.1 | 63.3 (±2.4) | 0.1 | 61.6 (±2.5) | 0.1 | 67.7 (±4.0) | 0.1 | 53.2 (±2.7) | 0.1 | 44.5 (±2.4) |

Adjusted mean intakes and standard errors (g or ml) exclude 1 person with missing age.

### Supplementary Table 3. Participant characteristics and daily energy and macronutrient intakes in 4,700 FEED participants

|  |  | | **Overall** | | | | **Omnivore** | | | | **Flexitarian** | | | | | **Pescatarian** | | | | **Vegetarian** | | | | **Vegan** | | | |  |
| --- | --- | --- | --- | --- | --- | --- | --- | --- | --- | --- | --- | --- | --- | --- | --- | --- | --- | --- | --- | --- | --- | --- | --- | --- | --- | --- | --- | --- |
|  | **N** | | 4,700 | | | | 1,216 | | | | 1,071 | | | | | 426 | | | | 899 | | | | 1,088 | | | | ***P*** ^‡^ |
|  | N% | | 100% | | | | 25.9% | | | | 22.8% | | | | | 9.1% | | | | 19.1% | | | | 23.1% | | | |  |
|  | **Characteristics** | | **N** | | **%** | | **N** | | **%** | | **N** | | | **%** | | **N** | | **%** | | **N** | | **%** | | **N** | | **%** | |  |
|  | Gender | |  | |  | |  | |  | |  | | |  | |  | |  | |  | |  | |  | |  | | <0.001 |
|  | Women | | 3,868 | | 82.3 | | 996 | | 81.9% | | 932 | | | 87.0% | | 372 | | 87.3% | | 749 | | 83.3% | | 820 | | 75.4% | |  |
|  | Men | | 770 | | 16.4 | | 214 | | 17.6% | | 128 | | | 12.0% | | 53 | | 12.4% | | 141 | | 15.7% | | 234 | | 21.5% | |  |
|  | Prefer not to say | | 61 | | 1.3 | | 6 | | 0.5% | | 11 | | | 1.0% | | - | | - | | 9 | | 1.0% | | 34 | | 3.1% | |  |
|  |  | | **Mean** | | **SD** | | **Mean** | | **SD** | | **Mean** | | | **SD** | | **Mean** | | **SD** | | **Mean** | | **SD** | | **Mean** | | **SD** | |  |
|  | Age (years) | | 52.3 | | 15.9 | | 53.6 | | 14.7 | | 52.7 | | | 16.9 | | 54.5 | | 16.1 | | 53.3 ^a^ | | 16.3 | | 48.9 | | 15.2 | | <0.001 |
|  | BMI (kg/m^2^) | | 24.4 | | 4.8 | | 25.6 | | 5.1 | | 23.7 | | | 4.3 | | 24.0 | | 4.7 | | 24.4 | | 4.9 | | 23.7 | | 4.4 | | <0.001 |
|  | Women | | 24.7 | | 4.1 | | 25.6 | | 5.3 | | 23.6 | | | 4.5 | | 24.0 | | 4.9 | | 24.2 | | 4.9 | | 23.7 | | 4.5 | |  |
|  | Men | | 24.3 | | 4.9 | | 25.8 | | 4.4 | | 24.0 | | | 3.1 | | 24.2 | | 3.3 | | 25.2 | | 5.0 | | 24.0 | | 3.5 | |  |
|  | Prefer not to say | | 23.6 | | 4.6 | | 24.0 | | 3.3 | | 24.1 | | | 4.7 | | - | | *-* | | 22.4 | | 3.0 | | 23.6 | | 5.2 | |  |
|  | **Mean nutrient Intakes**^†^ | | **Mean** | | **SD** | | **Mean** | | **SD** | | **Mean** | | | **SD** | | **Mean** | | **SD** | | **Mean** | | **SD** | | **Mean** | | **SD** | |  |
| Protein g/day | | 89.8 | | 29.0 | | 110.7 | | 35.0 | | 93.2 | | 21.2 | | | 88.3 | | 22.3 | | 76.4^a^ | | 19.5 | | 74.7^a^ | | 21.7 | |  | |
| Energy (kJ) | | 10024 | | 2084 | | 10217^b^ | | 2066 | | 10049^a,b^ | | 2036 | | | 10064^a,b^ | | 2054 | | 9828^a^ | | 2068 | | 9929^a^ | | 2157 | |  | |
| Energy (kcal) | | 2387 | | 497 | | 2435^b^ | | 485 | | 2392^b^ | | | 489 | | 2396^a,b^ | | 493 | | 2339^a^ | | 514 | | 2363^a^ | | 497 | |  | |
| Fat (%E) | | 36.8 | | 8.1 | | 39.8 | | 10.9 | | 36.3^a^ | | | 6.3 | | 35.6^a^ | | 7.1 | | 35.3^a^ | | 6.3 | | 35.7^a^ | | 6.9 | |  | |
| Carbohydrate (%E) | | 45.0 | | 9.8 | | 38.7 | | 13.2 | | 45.0^a^ | | | 6.7 | | 46.3^a^ | | 7.2 | | 48.6^b^ | | 6.6 | | 48.8^b^ | | 6.9 | |  | |
| Alcohol (%E) | | 2.7 | | 3.5 | | 2.9^a^ | | 4.0 | | 2.8^a^ | | | 3.2 | | 3.1^a^ | | 3.8 | | 2.7^a^ | | 3.3 | | 2.2 | | 3.1 | |  | |
| Protein (%E) | | 14.4 | | 3.6 | | 17.4 | | 4.3 | | 14.9 | | | 2.4 | | 14.1 | | 2.3 | | 12.4 | | 2.0 | | 12.0 | | 2.3 | |  | |
|  |  | | **N** | | **%** | | **N** | | **%** | | **N** | | | **%** | | **N** | | **%** | | **N** | | **%** | | **N** | | **%** | |  |
|  | **Proportion at or above protein EAR** | | 4,637 | | 98.7 | | 1215 | | 99.9%^c^ | | 1069 | | | 99.8%^c^ | | 424 | | 99.5%^bc^ | | 881 | | 98.0%^ab^ | | 1049 | | 96.4%^a^ | | <0.001 |

BMI = Body Mass Index; %E = Percent energy, EAR = Estimated Average Requirement

Excludes 1,413 participants with missing body weight data and EI:BMR <1.2 from total 6,113 analysis sample.

Age and mean nutrient intakes additionally exclude 1 participant with missing age.

a,b Estimates sharing a letter are not significantly different (p ≥ 0.05), based on pairwise comparisons of adjusted means following ANOVA with Bonferroni correction

All mean nutrient intakes P heterogeneity <0.001

^†^ Mean intakes are adjusted for age and gender

^‡^ P values for ANOVA and Chi-square tests

Data not shown where categories have <5 participants

### Supplementary Table 4. Mean daily intakes (g or ml) of all foods and beverages adjusted for age and gender and the respective protein contribution (%) by diet group in 4,700 FEED participants

| Food Item | Omnivore | | Flexitarian | | Pescatarian | | Vegetarian | | Vegan | |
| --- | --- | --- | --- | --- | --- | --- | --- | --- | --- | --- |
|  | Percentage of Total Protein (%) | Mean Intake (±SE) (g or ml) | Percentage of Total Protein (%) | Mean Intake (±SE) (g or ml) | Percentage of Total Protein (%) | Mean Intake (±SE) (g or ml) | Percentage of Total Protein (%) | Mean (±SE) Intake (g or ml) | Percentage of Total Protein (%) | Mean Intake (±SE) (g or ml) |
| All-Bran | 0.1 | 1.0 (±0.1) | 0.2 | 1.0 (±0.1) | 0.2 | 1.0 (±0.2) | 0.1 | 0.7 (±0.2) | 0.2 | 0.9 (±0.1) |
| Almond Milk | 0.0 | 7.5 (±1.6) | 0.1 | 10.1 (±1.7) | 0.1 | 12.4 (±2.7) | 0.1 | 12.0 (±1.9) | 0.1 | 13.2 (±1.7) |
| Apples, pears | 0.2 | 53.1 (±2.1) | 0.4 | 76.9 (±2.2) | 0.4 | 70.0 (±3.5) | 0.4 | 65.9 (±2.4) | 0.4 | 76.7 (±2.2) |
| Avocado | 0.4 | 24.0 (±1.1) | 0.6 | 28.2 (±1.2) | 0.8 | 36.5 (±1.9) | 0.6 | 25.8 (±1.3) | 0.8 | 32.2 (±1.2) |
| Bacon | 3.6 | 16.0 (±0.3) | 1.5 | 5.6 (±0.3) | 0.0 | 0.1 (±0.6) | 0.0 | 0.0 (±0.4) | 0.0 | 0.0 (±0.3) |
| Baked Beans | 0.6 | 13.3 (±0.6) | 0.7 | 14.3 (±0.7) | 1.0 | 19.0 (±1.0) | 1.3 | 20.3 (±0.7) | 1.7 | 25.3 (±0.7) |
| Banana | 0.4 | 33.9 (±1.3) | 0.6 | 46.7 (±1.4) | 0.7 | 48.7 (±2.2) | 0.7 | 46.5 (±1.5) | 0.9 | 59.1 (±1.4) |
| Beer, lager or cider | 0.1 | 47 (±3.3) | 0.1 | 49.7 (±3.5) | 0.1 | 62 (±5.6) | 0.2 | 58.2 (±3.8) | 0.2 | 49.6 (±3.5) |
| Biscuits | 0.3 | 5.3 (±0.3) | 0.4 | 6.2 (±0.3) | 0.4 | 5.7 (±0.5) | 0.5 | 7.1 (±0.3) | 0.4 | 5.2 (±0.3) |
| Boiled, mashed, or jacket potatoes | 0.9 | 48.5 (±1.3) | 1.0 | 47.4 (±1.4) | 1.1 | 47.6 (±2.2) | 1.4 | 53.8 (±1.5) | 1.3 | 49.6 (±1.4) |
| Bran flakes, Weetabix, muesli | 0.9 | 9.5 (±0.5) | 1.4 | 12.5 (±0.5) | 1.5 | 11.5 (±0.9) | 1.6 | 11.4 (±0.6) | 1.4 | 10.5 (±0.5) |
| Broccoli, spring greens, kale, spinach | 1.0 | 42.0 (±1.2) | 1.4 | 46.8 (±1.2) | 1.6 | 51.6 (±1.9) | 1.5 | 44.2 (±1.3) | 2.2 | 61.0 (±1.2) |
| Brown rice | 0.4 | 13.3 (±0.7) | 0.8 | 20.6 (±0.8) | 0.9 | 22 (±1.3) | 1.0 | 20.7 (±0.9) | 1.4 | 29.9 (±0.8) |
| Brown/granary bread or bread products | 1.9 | 20.6 (±1.1) | 2.7 | 25.8 (±1.1) | 3.0 | 27.3 (±1.8) | 3.5 | 26.1 (±1.2) | 3.4 | 26.1 (±1.1) |
| Butter | 0.0 | 8.3 (±0.2) | 0.0 | 5.1 (±0.2) | 0.0 | 4.2 (±0.3) | 0.0 | 4.0 (±0.2) | 0.0 | 0.3 (±0.2) |
| Cabbage, Brussels sprouts, Cauliflower, coleslaw | 0.5 | 27.1 (±0.8) | 0.6 | 27.8 (±0.8) | 0.7 | 29.1 (±1.3) | 0.8 | 27.1 (±0.9) | 1.0 | 36.7 (±0.8) |
| Cake | 0.8 | 16.0 (±0.6) | 1.1 | 18.2 (±0.6) | 1.0 | 14.6 (±0.9) | 1.3 | 17.6 (±0.7) | 0.7 | 12.2 (±0.6) |
| Carrot | 0.1 | 24.0 (±0.6) | 0.2 | 28 (±0.7) | 0.2 | 26.6 (±1.1) | 0.2 | 26.2 (±0.7) | 0.2 | 32.5 (±0.7) |
| Cereal Bar | 0.1 | 2.4 (±0.1) | 0.2 | 2.5 (±0.2) | 0.2 | 2.6 (±0.2) | 0.2 | 3.0 (±0.2) | 0.2 | 2.3 (±0.2) |
| Cheese | 3.7 | 22.1 (±0.5) | 3.7 | 18.2 (±0.5) | 3.9 | 17.8 (±0.8) | 5.2 | 21.0 (±0.6) | 0.0 | 0.0 (±0.5) |
| Cheese Sauce | 0.5 | 5.4 (±0.2) | 0.5 | 4.1 (±0.2) | 0.5 | 4.8 (±0.3) | 0.6 | 4.4 (±0.2) | 0.0 | 0.2 (±0.2) |
| Chickpeas | 0.5 | 8.7 (±0.6) | 1.3 | 16.2 (±0.6) | 1.8 | 22.1 (±1.0) | 1.9 | 19.4 (±0.7) | 3.0 | 30.8 (±0.6) |
| Chips, roast potatoes | 0.6 | 27.3 (±0.7) | 0.6 | 21.8 (±0.8) | 0.7 | 26.3 (±1.2) | 0.9 | 28.0 (±0.9) | 1.0 | 28.9 (±0.8) |
| Chocolate Spread | 0.0 | 0.8 (±0.1) | 0.0 | 0.9 (±0.1) | 0.0 | 0.8 (±0.1) | 0.1 | 1.0 (±0.1) | 0.0 | 0.7 (±0.1) |
| Chocolate or sweets, all types | 0.3 | 10.4 (±0.4) | 0.4 | 11.5 (±0.5) | 0.4 | 11.5 (±0.7) | 0.6 | 13.5 (±0.5) | 0.4 | 9.6 (±0.5) |
| Cholesterol-lowering margarine | 0.0 | 0.3 (±0.1) | 0.0 | 0.3 (±0.1) | 0.0 | 0.4 (±0.1) | 0.0 | 0.4 (±0.1) | 0.0 | 0.1 (±0.1) |
| Coconut Milk | 0.0 | 1.9 (±0.9) | 0.0 | 3.0 (±1.0) | 0.0 | 7.3 (±1.6) | 0.0 | 3.5 (±1.1) | 0.0 | 4.8 (±1.0) |
| Coffee, all types | 0.4 | 275.4 (±6.5) | 0.5 | 270.4 (±6.9) | 0.5 | 261.7 (±11.0) | 0.6 | 253.1 (±7.6) | 0.6 | 263.7 (±6.9) |
| Couscous, bulgur wheat, etc | 0.4 | 9.4 (±0.5) | 0.8 | 15.8 (±0.6) | 1.0 | 19.1 (±0.9) | 0.9 | 15.7 (±0.6) | 1.2 | 19.7 (±0.6) |
| Crackers | 0.2 | 2.1 (±0.1) | 0.2 | 2.3 (±0.1) | 0.3 | 2.4 (±0.2) | 0.3 | 2.5 (±0.1) | 0.2 | 1.9 (±0.1) |
| Cream | 0.2 | 7.2 (±0.3) | 0.1 | 2.2 (±0.3) | 0.1 | 1.9 (±0.4) | 0.1 | 1.7 (±0.3) | 0.0 | 0.1 (±0.3) |
| Crispbread | 0.1 | 1.3 (±0.1) | 0.2 | 2.1 (±0.1) | 0.2 | 1.9 (±0.2) | 0.2 | 1.7 (±0.1) | 0.2 | 1.6 (±0.1) |
| Crisps | 0.4 | 8.5 (±0.4) | 0.4 | 7.6 (±0.4) | 0.5 | 8.9 (±0.6) | 0.7 | 10.6 (±0.4) | 0.7 | 9.3 (±0.4) |
| Dairy full fat yogurt | 1.4 | 32.2 (±1.1) | 1.7 | 31.8 (±1.1) | 1.4 | 23.4 (±1.8) | 1.5 | 23.5 (±1.2) | 0.0 | 1.1 (±1.1) |
| Dairy ice cream | 0.3 | 12.6 (±0.4) | 0.3 | 10.5 (±0.4) | 0.3 | 8.7 (±0.7) | 0.4 | 9.5 (±0.5) | 0.0 | 0.4 (±0.4) |
| Dried fruit | 0.2 | 11.3 (±0.7) | 0.4 | 17.2 (±0.7) | 0.5 | 18.1 (±1.2) | 0.5 | 15.4 (±0.8) | 0.7 | 23.1 (±0.7) |
| Dried meats | 0.9 | 3.8 (±0.1) | 0.5 | 1.9 (±0.1) | 0.0 | 0.1 (±0.1) | 0.0 | 0.0 (±0.1) | 0.0 | 0.0 (±0.1) |
| Eggs | 5.7 | 48.2 (±1.3) | 4.5 | 32.6 (±1.5) | 5.4 | 40.9 (±2.3) | 5.4 | 35.6 (±1.7) | 0.2 | 0.9 (±1.4) |
| Falafel | 0.3 | 3.8 (±0.3) | 0.6 | 7.3 (±0.3) | 0.8 | 8.8 (±0.4) | 0.9 | 9.0 (±0.3) | 1.1 | 9.7 (±0.3) |
| Fish fried in batter/crumbs | 1.0 | 8.5 (±0.2) | 1.2 | 7.9 (±0.2) | 1.6 | 10.1 (±0.3) | 0.0 | 0.3 (±0.2) | 0.0 | 0.1 (±0.2) |
| Fizzy soft drinks | 0.0 | 20.2 (±1.7) | 0.0 | 11.5 (±1.8) | 0.0 | 14.4 (±2.8) | 0.0 | 15.7 (±1.9) | 0.0 | 11.4 (±1.8) |
| Fruit pie, fruit tart, crumble | 0.1 | 4.3 (±0.2) | 0.1 | 4.4 (±0.2) | 0.1 | 4.2 (±0.3) | 0.2 | 4.9 (±0.2) | 0.1 | 4.1 (±0.2) |
| Fruit squash or cordial | 0.0 | 112.8 (±5.6) | 0.0 | 70.4 (±6) | 0.0 | 77.4 (±9.5) | 0.0 | 84.3 (±6.5) | 0.0 | 73.4 (±6.0) |
| Goat/sheep's milk | 0.1 | 2.1 (±0.5) | 0.1 | 1.4 (±0.5) | 0.1 | 1.5 (±0.8) | 0.0 | 1.0 (±0.6) | 0.0 | 0.1 (±0.5) |
| Guacamole | 0.0 | 1.8 (±0.1) | 0.0 | 1.7 (±0.1) | 0.0 | 2.0 (±0.2) | 0.0 | 2.1 (±0.1) | 0.1 | 2.3 (±0.1) |
| Hemp Milk | 0.0 | 0.1 (±0.2) | 0.0 | 0.1 (±0.2) | 0.0 | 0.3 (±0.4) | 0.0 | 0.2 (±0.2) | 0.0 | 0.7 (±0.2) |
| Hot chocolate, milkshake | 0.1 | 16.6 (±1.1) | 0.1 | 16.3 (±1.2) | 0.2 | 17.0 (±1.9) | 0.2 | 18.8 (±1.3) | 0.2 | 16.1 (±1.2) |
| Huel shake/bar | 0.1 | 5.9 (±3.3) | 0.1 | 4.4 (±3.3) | 0.1 | 2.8 (±4.6) | 0.2 | 11.9 (±3.3) | 0.6 | 15.7 (±2.3) |
| Hummus | 0.3 | 3.8 (±0.4) | 0.6 | 7.1 (±0.4) | 0.7 | 8.7 (±0.6) | 0.9 | 8.9 (±0.4) | 1.5 | 14.7 (±0.4) |
| Jam, marmalade, honey | 0.0 | 5 (±0.2) | 0.0 | 5.9 (±0.3) | 0.0 | 5.5 (±0.4) | 0.0 | 5.6 (±0.3) | 0.0 | 4.5 (±0.3) |
| Lard/dripping | 0.0 | 1.5 (±0.1) | 0.0 | 0.1 (±0.1) | 0.0 | 0.0 (±0.1) | 0.0 | 0.0 (±0.1) | 0.0 | 0.0 (±0.1) |
| Low calorie soft drinks | 0.0 | 63.6 (±4.4) | 0.0 | 45.8 (±4.7) | 0.0 | 49.3 (±7.4) | 0.0 | 67.8 (±5.1) | 0.0 | 41.2 (±4.7) |
| Low-fat spread | 0.0 | 0.4 (±0.0) | 0.0 | 0.4 (±0.1) | 0.0 | 0.3 (±0.1) | 0.0 | 0.3 (±0.1) | 0.0 | 0.1 (±0.1) |
| Low-fat yogurt | 1.1 | 22.1 (±1.2) | 1.7 | 28.7 (±1.3) | 1.9 | 29.7 (±2.0) | 1.7 | 21.7 (±1.4) | 0.0 | 0.4 (±1.2) |
| Marmite | 0.3 | 1 (±0.1) | 0.5 | 1.3 (±0.1) | 0.6 | 1.4 (±0.2) | 0.9 | 2.0 (±0.1) | 1.2 | 2.6 (±0.1) |
| Mayonnaise | 0.1 | 6.3 (±0.2) | 0.1 | 4.5 (±0.2) | 0.1 | 4.6 (±0.3) | 0.0 | 3.3 (±0.2) | 0.0 | 0.1 (±0.2) |
| Meat Soup | 1.0 | 17.4 (±0.5) | 0.7 | 9.9 (±0.5) | 0.3 | 3.8 (±0.8) | 0.0 | 0.1 (±0.6) | 0.0 | 0.0 (±0.5) |
| Meat alternatives | 0.1 | 0.5 (±0.3) | 0.2 | 1.2 (±0.4) | 0.7 | 3.3 (±0.6) | 1.1 | 4.1 (±0.4) | 1.6 | 6.5 (±0.4) |
| Milk-based pudding | 0.3 | 9.8 (±0.4) | 0.3 | 8.7 (±0.4) | 0.3 | 8.2 (±0.6) | 0.4 | 9.5 (±0.4) | 0.2 | 5.0 (±0.4) |
| Milk-based smoothie | 0.2 | 8.6 (±1.0) | 0.3 | 9.6 (±1.1) | 0.3 | 10.4 (±1.8) | 0.3 | 8.6 (±1.2) | 0.1 | 13.3 (±1.1) |
| Mushrooms | 0.2 | 13.1 (±0.4) | 0.2 | 14.2 (±0.5) | 0.3 | 17 (±0.8) | 0.3 | 16.3 (±0.5) | 0.4 | 21.3 (±0.5) |
| Nut and/or dried fruit bar | 0.2 | 2.4 (±0.2) | 0.3 | 3.4 (±0.2) | 0.4 | 3.9 (±0.3) | 0.4 | 3.7 (±0.2) | 0.5 | 4.7 (±0.2) |
| Nut butters (with added sugar) | 0.1 | 0.6 (±0.1) | 0.1 | 0.6 (±0.1) | 0.2 | 0.7 (±0.2) | 0.3 | 0.9 (±0.1) | 0.4 | 1.3 (±0.1) |
| Nut butters (without sugar) | 0.6 | 3.3 (±0.3) | 1.2 | 5.5 (±0.3) | 1.4 | 6.3 (±0.5) | 1.5 | 5.8 (±0.4) | 2.8 | 10.3 (±0.3) |
| Nuts | 1.7 | 9.4 (±0.6) | 2.9 | 13.3 (±0.6) | 3.1 | 13.4 (±1.0) | 3.4 | 13.0 (±0.7) | 5.0 | 20.7 (±0.6) |
| Oat Milk | 0.1 | 16.6 (±3.4) | 0.4 | 50.2 (±3.6) | 0.5 | 69.7 (±5.8) | 0.6 | 70.9 (±4.0) | 1.0 | 118.1 (±3.6) |
| Oatcakes | 0.2 | 2.0 (±0.2) | 0.4 | 3.2 (±0.2) | 0.4 | 3.0 (±0.3) | 0.3 | 2.6 (±0.2) | 0.3 | 2.7 (±0.2) |
| Offal | 0.9 | 4.7 (±0.2) | 0.3 | 1.4 (±0.2) | 0.0 | 0.0 (±0.4) | 0.0 | 0.0 (±0.3) | 0.0 | 0 (±0.2) |
| Oily fish | 3.7 | 18.1 (±0.5) | 4.3 | 17.8 (±0.5) | 5.1 | 19.2 (±0.8) | 0.1 | 0.2 (±0.5) | 0.0 | 0.3 (±0.5) |
| Olive Oil | 0.0 | 6.4 (±0.2) | 0.0 | 8.1 (±0.2) | 0.0 | 6.9 (±0.3) | 0.0 | 6.5 (±0.2) | 0.0 | 6.4 (±0.2) |
| Onions, leeks | 0.6 | 39.5 (±1.0) | 0.8 | 42.8 (±1) | 0.9 | 45.1 (±1.6) | 1.1 | 44.7 (±1.1) | 1.2 | 52.7 (±1.0) |
| Oranges, satsumas, grapefruit | 0.3 | 39.1 (±1.6) | 0.4 | 49.1 (±1.8) | 0.4 | 44.7 (±2.8) | 0.4 | 43.6 (±1.9) | 0.5 | 53.7 (±1.7) |
| Other Milk (Dairy) | 0.2 | 6.8 (±1.0) | 0.2 | 6.1 (±1.1) | 0.0 | 1.1 (±1.7) | 0.1 | 2.1 (±1.1) | 0.0 | 0.2 (±1.0) |
| Other beans or lentils | 0.8 | 13.0 (±0.8) | 1.9 | 24.9 (±0.9) | 2.4 | 30.7 (±1.4) | 2.7 | 29.3 (±1.0) | 4.4 | 47.2 (±0.9) |
| Other bread products | 0.4 | 6.0 (±0.3) | 0.5 | 6.2 (±0.3) | 0.6 | 6.8 (±0.4) | 0.7 | 7.5 (±0.3) | 0.7 | 6.6 (±0.3) |
| Other breakfast cereal | 0.3 | 3.5 (±0.2) | 0.3 | 3.0 (±0.2) | 0.2 | 2.3 (±0.4) | 0.4 | 3.4 (±0.3) | 0.3 | 2.5 (±0.2) |
| Other fat spread/margarine | 0.0 | 0.5 (±0.1) | 0.0 | 0.6 (±0.1) | 0.0 | 0.3 (±0.1) | 0.0 | 0.3 (±0.1) | 0.0 | 0.1 (±0.1) |
| Other fresh fruit | 0.4 | 51.6 (±2.0) | 0.7 | 66.5 (±2.1) | 0.7 | 63.3 (±3.3) | 0.7 | 58.0 (±2.3) | 0.8 | 73.3 (±2.1) |
| Other meal replacement shake/bar | 0.1 | 10.4 (±2.1) | 0.2 | 7.6 (±2.1) | 0.1 | 4.1 (±2.9) | 0.1 | 4.3 (±2.1) | 0.2 | 1.3 (±1.5) |
| Other milk alternatives | 0.0 | 1.6 (±0.8) | 0.0 | 1.8 (±0.8) | 0.0 | 3.0 (±1.3) | 0.0 | 1.6 (±0.9) | 0.0 | 3.9 (±0.8) |
| Other processed meat | 1.0 | 7.6 (±0.2) | 0.6 | 3.7 (±0.2) | 0.0 | 0.3 (±0.3) | 0.0 | 0.1 (±0.2) | 0.0 | 0.0 (±0.2) |
| Other sauces, white sauce, gravy | 0.1 | 7.1 (±0.2) | 0.1 | 5.1 (±0.2) | 0.1 | 5.1 (±0.3) | 0.2 | 6.8 (±0.2) | 0.0 | 4.8 (±0.2) |
| Other vegetable oil | 0.0 | 2.6 (±0.1) | 0.0 | 2.8 (±0.1) | 0.0 | 2.9 (±0.2) | 0.0 | 3.3 (±0.1) | 0.0 | 3.7 (±0.1) |
| Other vegetables | 0.3 | 29.6 (±1.1) | 0.5 | 37.2 (±1.2) | 0.5 | 36.0 (±1.8) | 0.6 | 35.1 (±1.3) | 0.8 | 45.1 (±1.2) |
| Other veggie burgers or sausages | 0.2 | 2.1 (±0.3) | 0.9 | 5.9 (±0.3) | 1.5 | 9.2 (±0.4) | 2.1 | 11.6 (±0.3) | 2.1 | 10.6 (±0.3) |
| Pea Milk | 0.0 | 0.1 (±0.2) | 0.0 | 0.3 (±0.2) | 0.0 | 0.0 (±0.4) | 0.0 | 0.0 (±0.2) | 0.0 | 0.4 (±0.2) |
| Peas, green beans | 1.1 | 24.9 (±0.7) | 1.5 | 27.7 (±0.7) | 1.8 | 30.3 (±1.1) | 1.8 | 26.6 (±0.8) | 2.2 | 32.5 (±0.7) |
| Pizza | 1.3 | 12.5 (±0.5) | 1.7 | 12.8 (±0.6) | 1.9 | 14.1 (±0.9) | 2.5 | 16.2 (±0.6) | 2.9 | 18.5 (±0.6) |
| Polyunsaturated margarine | 0.0 | 0.4 (±0.1) | 0.0 | 0.3 (±0.1) | 0.0 | 0.4 (±0.1) | 0.0 | 0.4 (±0.1) | 0.0 | 0.3 (±0.1) |
| Porridge, Ready brek, Overnight Oats | 0.6 | 45.0 (±2.2) | 1.0 | 66.2 (±2.3) | 1.2 | 76.3 (±3.7) | 1.1 | 61.5 (±2.6) | 1.4 | 78.3 (±2.3) |
| Protein Bar (Hemp) | 0.0 | 0.0 (±0.0) | 0.0 | 0.0 (±0.0) | 0.0 | 0.0 (±0.0) | 0.0 | 0.0 (±0.0) | 0.0 | 0.0 (±0.0) |
| Protein Bar (Other) | 0.0 | 0.2 (±0.1) | 0.0 | 0.1 (±0.1) | 0.1 | 0.3 (±0.2) | 0.1 | 0.2 (±0.1) | 0.2 | 0.5 (±0.1) |
| Protein Bar (Pea) | 0.0 | 0.1 (±0.1) | 0.1 | 0.3 (±0.1) | 0.1 | 0.3 (±0.2) | 0.1 | 0.2 (±0.1) | 0.2 | 0.5 (±0.1) |
| Protein Bar (Peanut) | 0.0 | 0.0 (±0.0) | 0.0 | 0.1 (±0.0) | 0.0 | 0.1 (±0.0) | 0.0 | 0.0 (±0.0) | 0.0 | 0.0 (±0.0) |
| Protein Bar (Rice) | 0.0 | 0.0 (±0.0) | 0.0 | 0.0 (±0.0) | 0.0 | 0.0 (±0.0) | 0.0 | 0.0 (±0.0) | 0.0 | 0.0 (±0.0) |
| Protein Bar (Soy) | 0.0 | 0.0 (±0.1) | 0.0 | 0.1 (±0.1) | 0.1 | 0.3 (±0.1) | 0.1 | 0.2 (±0.1) | 0.2 | 0.4 (±0.1) |
| Protein Bar (Whey) | 0.1 | 0.5 (±0.1) | 0.2 | 0.5 (±0.1) | 0.1 | 0.2 (±0.1) | 0.1 | 0.2 (±0.1) | 0.0 | 0.0 (±0.1) |
| Protein Shake (Hemp) | 0.0 | 0.0 (±0.0) | 0.0 | 0.1 (±0.0) | 0.0 | 0.1 (±0.1) | 0.0 | 0.0 (±0.0) | 0.1 | 0.1 (±0.0) |
| Protein Shake (Other) | 0.1 | 0.2 (±0.1) | 0.1 | 0.2 (±0.1) | 0.2 | 0.3 (±0.2) | 0.3 | 0.4 (±0.1) | 0.6 | 0.6 (±0.1) |
| Protein Shake (Pea) | 0.1 | 0.1 (±0.1) | 0.3 | 0.4 (±0.1) | 0.4 | 0.5 (±0.2) | 0.4 | 0.5 (±0.1) | 1.4 | 1.0 (±0.1) |
| Protein Shake (Peanut) | 0.0 | 0.0 (±0.0) | 0.0 | 0.0 (±0.0) | 0.0 | 0.0 (±0.0) | 0.0 | 0.0 (±0.0) | 0.0 | 0.0 (±0.0) |
| Protein Shake (Rice) | 0.0 | 0.0 (±0.0) | 0.0 | 0.0 (±0.0) | 0.0 | 0.0 (±0.0) | 0.0 | 0.0 (±0.0) | 0.0 | 0.0 (±0.0) |
| Protein Shake (Soy) | 0.0 | 0.0 (±0.1) | 0.2 | 0.2 (±0.1) | 0.2 | 0.2 (±0.1) | 0.2 | 0.2 (±0.1) | 0.7 | 0.6 (±0.1) |
| Protein Shake (Whey) | 0.6 | 0.9 (±0.1) | 0.5 | 0.5 (±0.1) | 0.4 | 0.6 (±0.2) | 0.4 | 0.4 (±0.1) | 0.0 | 0.0 (±0.1) |
| Pure fruit juice or smoothie | 0.2 | 52.0 (±3.0) | 0.3 | 53.5 (±3.2) | 0.3 | 47.1 (±5.1) | 0.4 | 64.3 (±3.5) | 0.4 | 60.6 (±3.2) |
| Quorn | 0.2 | 2.0 (±0.4) | 0.8 | 5.8 (±0.4) | 1.9 | 13.0 (±0.6) | 3.0 | 17.4 (±0.4) | 1.3 | 6.9 (±0.4) |
| Rice Milk | 0.0 | 0.2 (±0.3) | 0.0 | 1.0 (±0.4) | 0.0 | 0.2 (±0.6) | 0.0 | 1.2 (±0.4) | 0.0 | 0.4 (±0.4) |
| Root vegetables | 0.2 | 14.9 (±0.5) | 0.2 | 18.2 (±0.6) | 0.2 | 18.2 (±0.9) | 0.3 | 17.4 (±0.6) | 0.3 | 22.1 (±0.6) |
| Salad cream, dressing, dips, low-fat mayo | 0.0 | 2.4 (±0.1) | 0.0 | 1.9 (±0.1) | 0.0 | 1.8 (±0.2) | 0.0 | 1.7 (±0.1) | 0.0 | 0.1 (±0.1) |
| Salad vegetables | 0.3 | 33.7 (±1.0) | 0.5 | 39.4 (±1.1) | 0.5 | 38.8 (±1.7) | 0.5 | 35.9 (±1.2) | 0.6 | 42.7 (±1.1) |
| Salted peanuts, other nuts or seeds | 0.6 | 3.4 (±0.2) | 0.8 | 3.9 (±0.2) | 0.9 | 4.4 (±0.4) | 1.0 | 4.3 (±0.3) | 1.2 | 5.1 (±0.2) |
| Sandwich slices | 0.7 | 3.6 (±0.1) | 0.3 | 1.2 (±0.1) | 0.0 | 0.0 (±0.1) | 0.0 | 0.0 (±0.1) | 0.0 | 0.0 (±0.1) |
| Sausages | 1.0 | 7.1 (±0.1) | 0.6 | 3.5 (±0.1) | 0.0 | 0.1 (±0.2) | 0.0 | 0.0 (±0.1) | 0.0 | 0.0 (±0.1) |
| Seitan | 0.0 | 0.2 (±0.2) | 0.2 | 0.7 (±0.2) | 0.3 | 1.5 (±0.3) | 0.5 | 2.1 (±0.2) | 1.5 | 5.7 (±0.2) |
| Semi Skimmed Milk | 3.6 | 113.9 (±3.5) | 3.6 | 97.2 (±3.7) | 3.2 | 77.8 (±5.9) | 3.0 | 63.9 (±4.0) | 0.0 | 2.6 (±3.7) |
| Shellfish | 0.9 | 5.0 (±0.2) | 0.9 | 4.1 (±0.2) | 1.1 | 4.6 (±0.3) | 0.0 | 0.1 (±0.2) | 0.0 | 0.0 (±0.2) |
| Skimmed Milk | 0.8 | 26 (±2.3) | 1.1 | 28.3 (±2.4) | 1.2 | 29.4 (±3.9) | 1.4 | 27.9 (±2.7) | 0.0 | 1.3 (±2.4) |
| Soy Milk | 0.2 | 7.5 (±3.2) | 0.6 | 18.4 (±3.4) | 1.0 | 30.7 (±5.4) | 1.7 | 44.9 (±3.7) | 4.3 | 107.3 (±3.4) |
| Soya burgers or sausages | 0.1 | 0.7 (±0.2) | 0.4 | 2.2 (±0.2) | 0.7 | 4.2 (±0.3) | 1.1 | 5.4 (±0.2) | 1.5 | 7.0 (±0.2) |
| Spirits | 0.0 | 3.7 (±0.2) | 0.0 | 3.1 (±0.2) | 0.0 | 3.5 (±0.3) | 0.0 | 3.4 (±0.2) | 0.0 | 2.6 (±0.2) |
| Sugar | 0.0 | 1.2 (±0.1) | 0.0 | 0.9 (±0.1) | 0.0 | 0.7 (±0.2) | 0.0 | 0.9 (±0.1) | 0.0 | 1.1 (±0.1) |
| Sugary cereal | 0.2 | 3.0 (±0.2) | 0.2 | 2.8 (±0.2) | 0.2 | 2.8 (±0.4) | 0.3 | 3.7 (±0.3) | 0.2 | 1.9 (±0.2) |
| Sweet peppers | 0.3 | 34.5 (±1.1) | 0.4 | 38.4 (±1.1) | 0.5 | 45.2 (±1.8) | 0.5 | 39.4 (±1.3) | 0.5 | 44.6 (±1.1) |
| Sweetcorn | 0.1 | 3.4 (±0.1) | 0.1 | 3.6 (±0.2) | 0.1 | 4.0 (±0.2) | 0.1 | 4.1 (±0.2) | 0.2 | 4.6 (±0.2) |
| Tahini | 0.2 | 1.3 (±0.2) | 0.5 | 2.3 (±0.2) | 0.5 | 2.6 (±0.3) | 0.7 | 3.0 (±0.2) | 1.1 | 4.9 (±0.2) |
| Tea, all types | 0.1 | 366.6 (±7.5) | 0.1 | 375 (±8.0) | 0.1 | 382.1 (±12.7) | 0.1 | 366.3 (±8.7) | 0.1 | 353.9 (±8.0) |
| Tempeh | 0.1 | 0.5 (±0.2) | 0.4 | 1.5 (±0.2) | 0.6 | 2.6 (±0.4) | 0.8 | 3.0 (±0.3) | 1.9 | 6.9 (±0.2) |
| Textured vegetable protein | 0.1 | 0.2 (±0.1) | 0.4 | 0.7 (±0.1) | 0.7 | 1.3 (±0.1) | 1.1 | 1.7 (±0.1) | 1.2 | 1.7 (±0.1) |
| Tinned or stewed fruit | 0.0 | 10.1 (±0.7) | 0.1 | 10.4 (±0.7) | 0.1 | 9.6 (±1.1) | 0.1 | 10.2 (±0.8) | 0.1 | 8.4 (±0.7) |
| Tofu | 0.2 | 2.5 (±0.5) | 0.7 | 7.4 (±0.5) | 1.0 | 11.0 (±0.8) | 1.3 | 12.2 (±0.6) | 2.8 | 24.7 (±0.5) |
| Tomato based sauce | 0.2 | 14.4 (±0.4) | 0.3 | 15.6 (±0.5) | 0.3 | 16.2 (±0.8) | 0.4 | 17.9 (±0.5) | 0.4 | 17.5 (±0.5) |
| Tomatoes | 0.4 | 51.5 (±1.3) | 0.5 | 58.7 (±1.4) | 0.6 | 56.9 (±2.2) | 0.7 | 59.4 (±1.5) | 0.7 | 64.1 (±1.4) |
| Tuna | 2.7 | 10 (±0.2) | 3.0 | 9.6 (±0.2) | 3.5 | 10.3 (±0.4) | 0.1 | 0.2 (±0.3) | 0.0 | 0.0 (±0.2) |
| Unprocessed beef | 8.6 | 34.3 (±0.9) | 2.2 | 7.6 (±0.9) | 0.0 | 0.3 (±1.5) | 0.0 | 0.1 (±1.0) | 0.0 | 0.0 (±0.9) |
| Unprocessed chicken, turkey | 8.7 | 33.3 (±0.5) | 4.6 | 15 (±0.5) | 0.2 | 0.5 (±0.8) | 0.0 | 0.1 (±0.5) | 0.0 | 0.0 (±0.5) |
| Unprocessed lamb | 3.4 | 13.8 (±0.4) | 1.4 | 4.8 (±0.4) | 0.0 | 0 .0 (±0.6) | 0.0 | 0.0 (±0.4) | 0.0 | 0.0 (±0.4) |
| Unprocessed pork | 4.4 | 17.4 (±0.4) | 1.4 | 5.1 (±0.4) | 0.0 | 0.0 (±0.7) | 0.0 | 0 (±0.5) | 0.0 | 0.0 (±0.4) |
| Vegan cheese | 0.0 | 0.5 (±0.2) | 0.0 | 1.0 (±0.2) | 0.1 | 1.9 (±0.3) | 0.1 | 2.8 (±0.2) | 0.4 | 8.9 (±0.2) |
| Vegan cream | 0.0 | 0.2 (±0.1) | 0.0 | 0.6 (±0.1) | 0.0 | 0.8 (±0.1) | 0.0 | 1.1 (±0.1) | 0.0 | 2.2 (±0.1) |
| Vegan fat spread/margarine | 0.1 | 2.7 (±0.9) | 0.2 | 7.9 (±1.0) | 0.3 | 9.8 (±1.6) | 0.4 | 14.9 (±1.1) | 1.2 | 39.6 (±1.0) |
| Vegan ice cream | 0.0 | 1.6 (±0.4) | 0.0 | 3.2 (±0.4) | 0.1 | 5.4 (±0.6) | 0.1 | 6.3 (±0.4) | 0.2 | 13.3 (±0.4) |
| Vegan mayonnaise | 0.0 | 0.4 (±0.1) | 0.0 | 0.9 (±0.1) | 0.0 | 1.7 (±0.2) | 0.0 | 2.6 (±0.1) | 0.0 | 5.5 (±0.1) |
| Vegan salad cream, dressing, dips, low-fat mayo | 0.0 | 0.1 (±0.1) | 0.0 | 0.2 (±0.1) | 0.0 | 0.6 (±0.1) | 0.0 | 1.0 (±0.1) | 0.0 | 1.9 (±0.1) |
| Vegan salad cream, dressing, dips, low-fat mayo | 0.0 | 0.2 (±0.1) | 0.0 | 0.8 (±0.2) | 0.0 | 1.1 (±0.2) | 0.0 | 2.8 (±0.2) | 0.0 | 5.8 (±0.2) |
| Vegan yogurt | 0.0 | 647.9 (±9.3) | 0.0 | 702.4 (±10.0) | 0.0 | 661.7 (±15.8) | 0.0 | 618.8 (±10.9) | 0.0 | 665.3 (±9.9) |
| Vegetable soup | 0.6 | 32.1 (±1.4) | 1.0 | 45.5 (±1.5) | 1.1 | 43.8 (±2.3) | 1.4 | 48.3 (±1.6) | 1.4 | 50.7 (±1.5) |
| Water | 1.4 | 18.7 (±0.9) | 1.2 | 13.1 (±0.9) | 1.2 | 12.1 (±1.5) | 1.9 | 16.9 (±1.0) | 1.7 | 14.2 (±0.9) |
| White bread or bread products | 0.0 | 0.6 (±0.0) | 0.0 | 0.3 (±0.0) | 0.0 | 0.4 (±0.1) | 0.0 | 0.4 (±0.1) | 0.0 | 0.6 (±0.0) |
| White hard vegetable fat | 2.3 | 26.7 (±1.3) | 3.5 | 36.2 (±1.4) | 4.0 | 36.1 (±2.2) | 5.1 | 41.5 (±1.5) | 5.4 | 45.7 (±1.4) |
| White pasta/noodles | 1.6 | 36.1 (±1.2) | 2.1 | 39.3 (±1.3) | 2.2 | 39.7 (±2.0) | 2.8 | 41.9 (±1.4) | 2.5 | 34.2 (±1.2) |
| White rice | 0.7 | 27.2 (±0.8) | 0.8 | 25.2 (±0.9) | 0.8 | 24.5 (±1.4) | 0.9 | 25.0 (±1.0) | 1.0 | 23.8 (±0.9) |
| Whole Milk | 1.9 | 58.9 (±2.5) | 0.8 | 22.4 (±2.7) | 0.7 | 19.3 (±4.3) | 0.6 | 13.7 (±3.0) | 0.0 | 0.0 (±2.7) |
| Wholemeal bread or bread products | 2.9 | 13.4 (±0.3) | 3.2 | 12.3 (±0.3) | 3.7 | 12.9 (±0.5) | 0.1 | 0.2 (±0.4) | 0.0 | 0.2 (±0.3) |
| Wholemeal pasta | 0.7 | 16.6 (±1.0) | 1.5 | 26.9 (±1.1) | 1.7 | 29.9 (±1.7) | 1.9 | 30.0 (±1.2) | 2.6 | 39.0 (±1.1) |
| Wine, champagne, sherry | 0.1 | 70.3 (±2.8) | 0.1 | 67.4 (±3.0) | 0.1 | 72.0 (±4.8) | 0.1 | 57.4 (±3.3) | 0.1 | 48.8 (±3.0) |

Excludes 1,413 participants with missing body weight data and EI:BMR <1.2 from total 6,113 analysis sample.

Adjusted mean intakes and standard errors (g or ml) additionally exclude 1 person with missing age.
